# Heterogeneity of Insulin Resistance Surrogates in Thousands of Non-Diabetic Adults: Multi-Modal Data Reveals Discordant Metabolic Phenotypes

**DOI:** 10.64898/2026.05.02.26352290

**Authors:** Smadar Shilo, Yeela Talmor-Barkan, Maria Gorodetski, Dana Azouri, Anastasia Godneva, Eran Segal, Hagai Rossman

## Abstract

The transition from metabolic health to type 2 diabetes unfolds through progressive insulin resistance (IR), yet the gold-standard hyperinsulinemic-euglycemic clamp is inapplicable at population scale and fasting insulin is not uniformly available. Several surrogate measures have been described in the literature, but whether these surrogates identify the same individuals, and whether continuous glucose monitoring (CGM) or NMR metabolomics carry information beyond conventional markers, remains unresolved. Here, we analyzed IR surrogates in 10,114 non-diabetic adults (35-75 y) from the Human Phenotype Project (HPP), integrated with 14-day CGM, dual x-ray absorptiometry (DEXA) body composition, liver and carotid ultrasound, sleep monitoring, and NMR metabolomics and established sex-specific, age-resolved reference ranges. IR surrogates were moderately inter-correlated but captured distinct metabolic facets. We next focused on DEXA-derived visceral adipose tissue (VAT), one of the strongest correlates of clamp-measured insulin resistance. Our analysis showed that VAT can be reliably predicted from anthropometric measurements alone (R² = 0.659). However, it is only modestly predicted by CGM features alone (R^2^ = 0.078). Among CGM-derived features, markers of glycemic variability were stronger predictors of VAT than conventional mean-glucose metrics. Residual-based analyses identified individuals whose visceral adiposity was substantially higher than expected given their BMI or HbA1c levels. Notably, 1.2% of adults in the HPP cohort exhibited elevated visceral adiposity despite having both a normal BMI (< 25 kg/m²) and normoglycemic HbA1c (< 5.7%). These discordant subpopulations harbored adverse profiles across lipid, hepatic, vascular, sleep, and metabolomic domains. NMR lipoprotein subfractions (VLDL, HDL) discriminated discordant phenotypes. A CGM variability-only model separated discordant individuals at AUC = 0.63, with negligible gain from adding mean glucose. Findings were validated in an independent cohort with available fasting insulin data. Together, these results establish normative IR surrogate reference ranges, quantify the fraction of metabolically at-risk individuals missed by conventional BMI and HbA1c screening, and highlight CGM variability metrics and NMR lipoprotein profiling as complementary tools for early metabolic risk stratification.

**Graphical Abstract:** 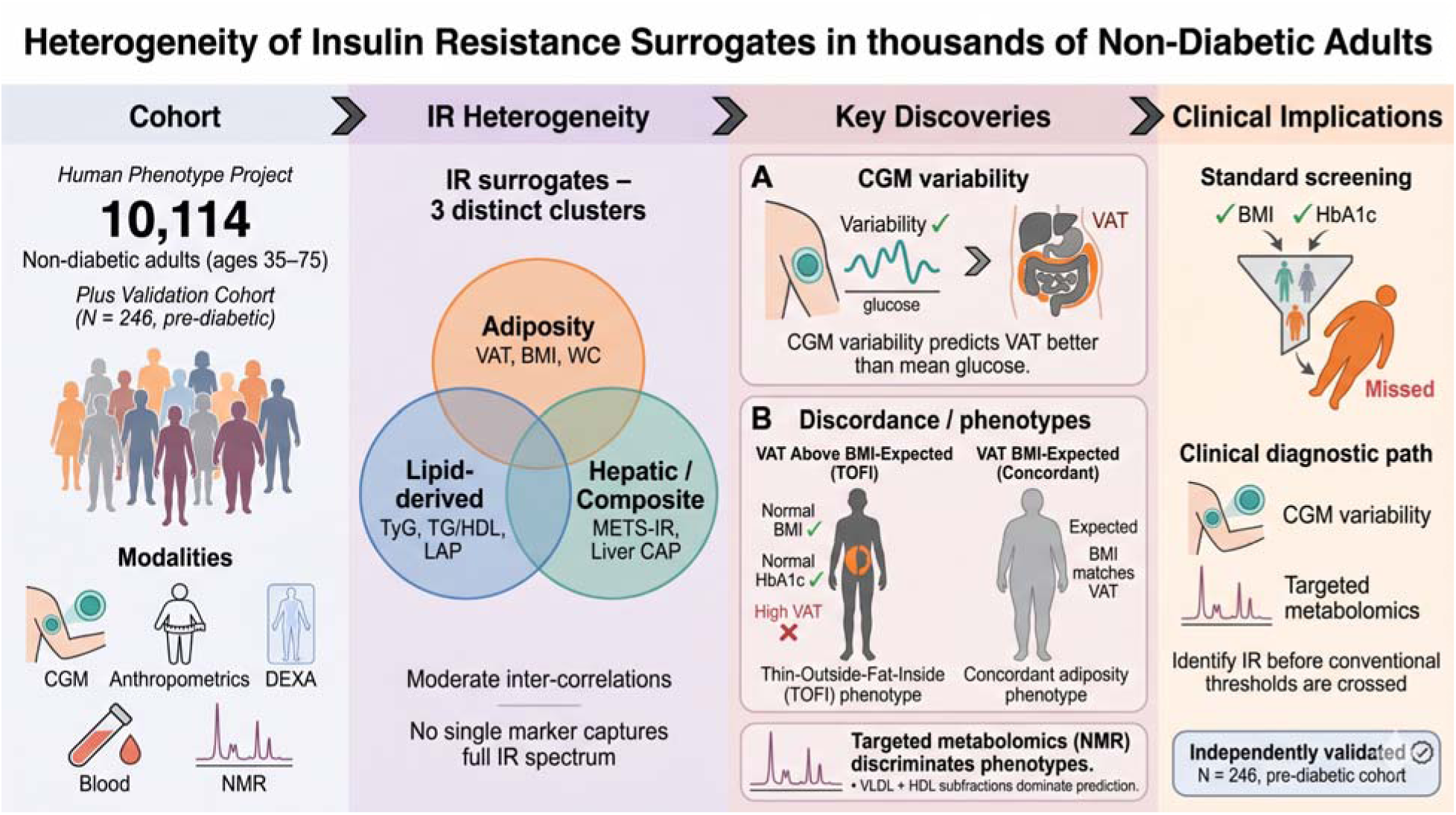

## Introduction

The transition from metabolic health to overt type 2 diabetes is rarely abrupt. Progressive insulin resistance (IR), defined as impaired insulin-mediated glucose uptake in skeletal muscle, impaired suppression of hepatic glucose production, and failure to restrain adipose tissue lipolysis usually progresses gradually over years to decades, driving vascular, hepatic, and adipose remodeling long before any diagnostic threshold is crossed ^1,2^. IR is considered as the primary pathophysiological process underlying the metabolic syndrome, and its early identification in non-diabetic individuals represents a window for preventive medicine strategies ^2^.

The hyperinsulinemic-euglycemic clamp has remained the reference standard for insulin sensitivity measurement for nearly five decades ^3^ but requires skilled personnel, continuous intravenous access and substantial cost, rendering it inapplicable for routine clinical care or population-scale research. In its absence, a range of validated surrogate measures have been developed to approximate clamp-derived IR from accessible clinical data, spanning anthropometric indices, fasting biochemistry, imaging-derived body composition, and hepatic markers, each correlating meaningfully with clamp-measured insulin sensitivity, though with varying accuracy and across distinct physiological domains ^4–7^.

IR is a multi-organ condition whose different facets are not equivalently captured by any single surrogate. Hepatic, skeletal muscle, and adipose tissue insulin sensitivity can be independently impaired ^2,8^, and surrogates designed to reflect one facet may systematically miss another. More specifically, adiposity-based surrogates capture the quantity and distribution of fat driving hepatic and peripheral IR ^5,9^; lipid-derived indices reflect the dyslipidemic phenotype arising from impaired lipolysis suppression ^9^ and hepatic markers index the downstream consequences of portal free fatty acid flux and hepatic steatosis ^10,11^. The extent to which IR surrogates agree on individual-level classification, and what fraction of non-diabetic adults harbor discordant metabolic risk profiles undetectable by conventional markers, has not been systematically established in a large deeply phenotyped cohort.

Currently, conventional clinical practice relies primarily on two markers for metabolic risk stratification: body mass index (BMI) and hemoglobin A1c (HbA1c). Both are practical and widely available, but neither was designed as a direct measure of IR. BMI does not distinguish visceral from subcutaneous adiposity and thereby individuals with normal BMI may harbor substantial visceral fat and IR, previously termed metabolically obese, normal-weight phenotype (MONW) ^11^ and more recently reframed as “thin outside, fat inside” (TOFI) to emphasize the role of hidden visceral and ectopic fat accumulation as the structural basis of this metabolic vulnerability ^12^. On the contrary, some obese individuals remain metabolically healthy ^13^. HbA1c reflects average glycemia over approximately three months ^14^ but is insensitive to the tissue-specific, dynamic aspects of IR that precede chronic hyperglycemia and may also reflect beta-cell dysfunction. Individuals whose visceral adiposity diverges from both BMI and HbA1c, classified as low-risk by conventional criteria yet harboring a metabolically adverse profile, represent a population that current screening systematically fails to detect, whose prevalence and characteristics in non-diabetic adults remain poorly defined. Residual-based approaches, in which visceral adiposity is regressed on BMI or HbA1c to quantify the discrepancy between observed and expected fat burden, offer a statistically principled framework for identifying such individuals ^15^.

Here, we address these questions using multi-modal deep phenotyping of 10,114 non-diabetic adults in the Human Phenotype Project (HPP) ^16^. We establish normative distributions and sex- and age-specific reference values for validated IR surrogates, characterize their inter-relationships and clinical domain associations. We next focused on Dual Energy X-ray Absorptiometry (DEXA)-measured visceral adipose tissue (VAT) mass, among the surrogates most strongly correlated with clamp-measured IR in the existing literature ^17^, and show it can be accurately estimated from routine clinical data, improving further with addition of waist and hip circumference measurements. We show that among Continuous glucose monitoring (CGM) derived features, glucose variability metrics, outperformed glucose level metrics in predicting VAT. We also identify a clinically important subset of individuals with normal BMI or HbA1c who harbor elevated VAT and a metabolically adverse profile across lipid, hepatic, and vascular domains. Finally we analyse whether NMR metabolomic profiles and CGM metrics can discriminate these discordant phenotypes.

## Methods

### Study Design and Participants

We analyzed participants from the HPP ^16,18^, a prospective deep phenotyping study conducted in Israel. Participants were adults aged 35-75 years. Each participant was connected to a blinded FreeStyle Libre Pro Flash continuous glucose monitor (FSL-CGM), which captured interstitial glucose readings at 15-minute intervals across 14 days. Throughout the monitoring period, all food and beverage consumption was recorded in real time, and sleep was assessed over three successive nights ^19^. Participants also completed a comprehensive set of clinical and laboratory evaluations, blood tests, anthropometric and vital sign assessments, hepatic ultrasonography, and body composition analysis using DEXA measured with the GE Lunar Prodigy Advance device by GE Healthcare, USA, and analyzed using the GE CoreScan and OneScan software applications. Blood tests were obtained by the participants’ Health Maintenance Organization (HMO), and were restricted to tests conducted within one year from the baseline visit. A complete description of inclusion and exclusion criteria, baseline and follow-up measurements, data acquisition procedures, and instrumentation is provided in prior publications detailing the HPP cohort ^16,18^.

Participants were excluded in a stepwise manner (Figure S1) based on the following criteria: insufficient CGM data quality, HbA1c levels of 6.5% or above, a documented diabetes diagnosis identified through ICD-11 codes (5A10, 5A11, 5A12, 5A14, or 5A40), or current use of antidiabetic medications (ATC code A10). After applying these filters, 10,114 participants remained in the final analytic sample.

External validation was performed in an independent cohort of 246 participants from the Personalized Postprandial Glucose Response-Targeting Diet (PPT) clinical trial ^20^, which enrolled pre-diabetic adults (HbA1c 5.7-6.5%). PPT included blinded CGM data collected using the FreeStyle Libre device at 15-minute intervals, and fasting insulin measurements enabling HOMA-IR computation (N = 165 with fasting insulin data). To compare CGM-IR associations, we constructed a 1:1 matched subcohort using caliper-based nearest-neighbor matching without replacement. Matching was exact on sex; standardized Euclidean distance on age, BMI, and HbA1c was thresholded at a caliper of 0.2 SD. PPT participants whose nearest HPP neighbor fell outside the caliper were dropped, yielding 135 matched PPT participants and 135 matched HPP participants. After matching, the two cohorts were well balanced on all three matching variables: age (52.0 vs. 52.0 years, p = 0.99), BMI (29.3 vs. 29.4 kg/m², p = 0.88), and HbA1c (5.8 vs. 5.8%, p = 0.72). Waist circumference and systolic blood pressure were also comparable between groups (p > 0.05).

### Measurements

CGM-derived measures were computed using the IGLU (Interpreting blood GLUcose data) package from pre-processed glucose time series after trimming the first and last recording days to avoid partial-day artifacts. Quality control confirmed all glucose values within the 40-300 mg/dL physiological range. Twenty-nine IGLU-derived metrics available in both HPP and PPT cohorts were used for all analyses, including mean, median, time-in-range(TIR)- TIR 70-180 mg/dl, TIR 63-140 mg/dl, time above 140 mg/dl, time above 180 mg/dl, time below 70 mg/dl; HbA1c estimates- Glucose management indicator (GMI), eA1c); variability measures: Standard deviation of all glucose values (SD), Coefficient of variation (CV), Mean Amplitude of Glycemic Excursions (MAGE), Mean difference between glucose values obtained at the same time of day (MODD), Mean Absolute Glucose (MAG), SD of rate-of-change, Continuous Overall Net Glycemic Action (CONGA), Glucose Variability Percentage (GVP); risk indices-Low Blood Glucose Index (LBGI), high Blood Glucose Index (HBGI), Average daily risk range (ADRR), hypo/hyper index, Index of Glycemic Control (IGC), and composite measures (J-index, Glycaemic Risk Assessment Diabetes Equation (GRADE), GRADE subcomponents, AUC). A description of the full list of CGM-derived measures is detailed in ^21^.

IR surrogate measures were obtained or computed from available HPP data and ranked by their published correlation with the hyperinsulinemic-euglycemic clamp (Table 1). These encompass adiposity-based measures (DEXA VAT mass, waist circumference, LAP index, android/gynoid ratio, BMI), lipid-derived composites (METS-IR, TyG index, TyG-BMI, TG/HDL ratio), a hepatic measure (liver attenuation coefficient), and glycemic measures (fasting glucose, HbA1c). ^22^. Computed formulae: METS-IR = ln(2 x FPG + TG) x BMI / ln(HDL); TyG index = ln(TG x FPG / 2); TG/HDL ratio = triglycerides / HDL cholesterol; LAP = [WC (cm) - 65] x TG (mmol/L) for men and [WC (cm) - 58] x TG (mmol/L) for women. HOMA-IR^23^, the most widely used clinical IR index, required fasting insulin and was available only in the PPT cohort used for validation.

**Table 1.** Insulin resistance surrogate measures, derivation formulae, and reference values obtained from the HPP. Abbreviations: A/G — android-to-gynoid; AUC-ROC — area under the receiver operating characteristic curve; BMI — body mass index; CAP — controlled attenuation parameter; DEXA — dual-energy X-ray absorptiometry; EHC — euglycemic-hyperinsulinemic clamp; FPG — fasting plasma glucose; HDL-C — high-density lipoprotein cholesterol; HOMA-IR — homeostatic model assessment of insulin resistance; IR — insulin resistance; LAP — lipid accumulation product; M-value — glucose infusion rate at steady-state clamp; METS-IR — metabolic score for insulin resistance; TG — fasting triglycerides; TG/HDL — triglyceride-to-HDL cholesterol ratio; TyG — triglyceride-glucose; VAT — visceral adipose tissue; WC — waist circumference

| Surrogate Measure | Derivation Formula | Evidence Summary | Male mean (SD), [95% CI] | Female mean (SD), [95% CI] |
| --- | --- | --- | --- | --- |
| Homeostatic Model Assessment of IR (HOMA-IR) | $(\text{Fasting insulin } [\mu\text{U/mL}] \times \text{Fasting glucose } [\text{mmol/L}]) / 22.5$; equivalent using mg/dL: $(\text{insulin } \times \text{glucose}) / 405$ | Bonora et al.<sup>7</sup> compared HOMA-IR against a 4-h EHC in 115 subjects: overall $r = -0.820$, non-diabetic subgroup $r = -0.754$. Meta-analysis<sup>24</sup> yielded a pooled $r = -0.60$ for log(HOMA-IR). | HOMA-IR was not available in the HPP cohort. | |
| DEXA Visceral Adipose Tissue Mass (VAT Mass) | DEXA-estimated VAT mass (g) | Abate et al.<sup>25</sup> established using CT-measured visceral fat and EHC that intra-abdominal fat was the strongest adiposity-based predictor of insulin-mediated glucose disposal ($r \approx -0.76$ with M-value). DEXA-derived VAT was shown to correlate with EHC-measured insulin sensitivity in children and adults<sup>26,27</sup>; formal Pearson $r$ vs. EHC M-value in non-diabetic adults estimated at $r \approx -0.65$–$-0.75$, extrapolated from CT-VAT data. | 1153.89 (703.74), [1129.65, 1178.12] | 599.55 (419.55), [585.75, 613.35] |
| Triglyceride-Glucose Index (TyG Index) | $\text{Ln}[(\text{Fasting triglycerides } [\text{mg/dL}] \times \text{Fasting glucose } [\text{mg/dL}]) / 2]$ | Guerrero -Romero et al.<sup>6</sup> directly validated against the EHC in 99 subjects: Pearson $r = -0.681$ with M-rates (non-diabetic $r = -0.670$, diabetic $r = -0.690$). Vasques et al.<sup>28</sup> independently confirmed TyG outperformed HOMA-IR. | 8.50 (0.51), [8.48, 8.52] | 8.29 (0.48), [8.27, 8.31] |
| Metabolic Score for Insulin Resistance (METS-IR) | $\text{Ln}[(2 \times \text{Fasting glucose } [\text{mg/dL}] + \text{Fasting triglycerides } [\text{mg/dL}]) \times \text{BMI } (\text{kg/m}^2) / \text{Ln}[\text{HDL-C } (\text{mg/dL})]$ | Bello-Chavolla et al.<sup>4</sup> validated against the EHC M-value adjusted for fat-free mass in 125 subjects (57 non-diabetic, 68 diabetic): Spearman $\rho = -0.622$ (adjusted for age and sex). In the original Mexican population, METS-IR outperformed TyG, TG/HDL-C, and TyG-BMI. | 39.37 (7.33), [39.08, 39.67] | 35.26 (7.47), [34.98, 35.55] |
| TyG-BMI Index (TyG-BMI) | $\text{Ln}[(\text{Fasting triglycerides } [\text{mg/dL}] \times \text{Fasting glucose } [\text{mg/dL}] / 2) \times \text{BMI } (\text{kg/m}^2)]$ | No direct EHC validation available. Er et al.<sup>29</sup> in 517 non-diabetic adults showed TyG-BMI outperformed TyG, TG/HDL-C, VAI, and LAP in identifying IR by HOMA-IR (AUC $\approx 0.84$–$0.88$). Consistently outperforms TyG alone in non-obese individuals, women, and younger adults. | 226.08 (38.28), [224.57, 227.60] | 211.84 (42.87), [210.20, 213.47] |
| Triglyceride to HDL Cholesterol Ratio (TG/HDL Ratio) | $\text{Fasting triglycerides } [\text{mg/dL}] / \text{HDL cholesterol } [\text{mg/dL}]$ | McLaughlin et al. <sup>30</sup> validated TG/HDL in 258 overweight non-diabetic subjects against steady-state plasma glucose (SSPG — insulin suppression test, not a standard EHC): TG/HDL $\geq 3.0$ identified IR with AUC-ROC = 0.781. A formal Pearson $r$ vs. EHC M-value was not reported. | 2.67 (2.02), [2.59, 2.74] | 1.76 (1.24), [1.72, 1.81] |
| Liver Attenuation | Ultrasound controlled attenuation parameter (dB/cm/MHz) | Seppälä-Lindroos et al. <sup>31</sup> used <sup>1</sup>H-MRS to measure liver fat and EHC + [<sup>3</sup>H]-glucose tracer to quantify hepatic glucose output in 30 healthy non-diabetic men: liver fat was independently related to impaired suppression of endogenous hepatic glucose production. Correlations with HOMA-IR were also shown, e.g. Li et al. <sup>32</sup>, showing r - 0.568. | 0.41 (0.10), [0.41, 0.41] | 0.40 (0.10), [0.40, 0.41] |
| Fasting Plasma Glucose (FPG) | Direct fasting venous glucose measurement (mg/dL or mmol/L) | In Bonora et al. <sup>7</sup>, FPG added little incremental predictive value beyond fasting insulin for clamp insulin sensitivity. Discriminative ability in a non-diabetic cohort with narrow FPG range is very limited. Included because it is universally available and anchors HOMA-IR and TyG. | 93.33 (9.06), [93.00, 93.66] | 91.01 (9.11), [90.70, 91.32] |
| Android to Gynoid Fat Ratio (A/G Ratio) | DEXA android region fat mass (g) / DEXA gynoid region fat mass (g) | No direct EHC validation. Bantle et al. <sup>26</sup> found A/G ratio to be the strongest DXA-derived predictor of IR in women (r - 0.52), outperforming total VAT, android fat mass, and BMI after adjustment. | 1.24 (0.25), [1.23, 1.25] | 0.90 (0.18), [0.90, 0.91] |
| Waist Circumference (WC) | Direct measurement at umbilical level (cm); | Pouliot et al. <sup>33</sup> established WC as a clinically relevant adiposity-based IR indicator. validated WC against both HOMA-IR and the euglycemic clamp <sup>34</sup>. | 93.62 (10.44), [93.32, 93.92] | 83.39 (10.91), [83.10, 83.68] |
| Body Mass Index (BMI) | Weight (kg) / Height<sup>2</sup> (m<sup>2</sup>) | Bogardus et al. <sup>35</sup> reported r ≈ -0.40 with EHC-measured insulin action. Cannot distinguish fat from lean mass or visceral from subcutaneous fat; | 26.39 (3.65), [26.29, 26.50] | 25.34 (4.28), [25.23, 25.46] |
| Lipid Accumulation Product (LAP) | Men: [WC (cm) - 65] × Fasting TG (mmol/L);<br>Women: [WC (cm) - 58] × Fasting TG (mmol/L) | Significant negative correlation was observed for the M value with LAP index (r=0.39). The LAP index showed higher predictive accuracy for the risk of insulin resistance as compared with HOMA-IR, QUICKI and FG-IR in non-obese, normoglycemic Asian Indian males <sup>22</sup>. | 41.33 (33.02), [40.06, 42.59] | 31.02 (26.30), [30.06, 31.99] |

NMR metabolomics data were obtained from the Nightingale Health platform (∼250 features including lipoprotein subfractions, amino acids, fatty acids, and glycoprotein acetyls). Features with >= 80% data completeness were retained.

### Statistical Analysis

We computed Spearman rank correlations with bootstrap 95% confidence intervals (1,000 iterations) between representative IR surrogates (DEXA VAT mass, METS-IR, TyG index, liver attenuation, TG/HDL ratio, waist circumference, A/G ratio) and clinical variables across six domains: body composition (total fat mass, android fat %), lipids (triglycerides, high-density lipoprotein (HDL), low-density lipoprotein (LDL), total cholesterol), vascular (systolic blood pressure (BP), diastolic BP, carotid intima-media thickness (IMT), estimated glomerular filtration rate (eGFR), retinal vessel width, fractal dimension, vessel density), hepatic (alanine aminotransferase (ALT), aspartate aminotransferase (AST), gamma-glutamyl transferase (GGT), liver elasticity, viscosity, speed of sound), sleep (oxygen desaturation index (ODI), apnea-hypopnea index (AHI), respiratory disturbance index (RDI), sleep efficiency, mean peripheral oxygen saturation (SpO2)), and dietary (calories, macronutrient percentages). P-values were corrected for multiple comparisons using the Benjamini-Hochberg false discovery rate procedure.

We conducted predictive modeling by Gradient boosting regression ^36^ (GBR; 200 estimators, maximum depth 4, learning rate 0.1) and linear regression were trained to predict DEXA VAT mass using 5-fold cross-validation (N = 6,799 participants with both DEXA and clinical data). The CGM-only model used 29 IGLU features, sex and age. The anthropometric model used sex, age, blood pressure, BMI, weight, and height. The circumference model added waist circumference, hip circumference, and waist-to-hip ratio. Feature importance was assessed via SHapley Additive exPlanations (SHAP) ^37^ framework.

Individuals with discordant IR status in whom VAT-based IR classification diverges from conventional clinical markers were identified by employing a residual-based approach ^15^. For each participant with available DEXA data, VAT mass was regressed on BMI (and separately on HbA1c) using sex-specific linear regression. The standardized residual captures how much an individual’s visceral adiposity deviates from the population-expected level given their BMI (or HbA1c). On the VAT-BMI axis, individuals with residuals exceeding +1 SD were classified as “VAT Above BMI-Expected” (higher visceral adiposity than their BMI predicts, capturing the TOFI phenotype^12^), those below -1 SD as “VAT Below BMI-Expected” (lower visceral adiposity than expected, analogous to the metabolically healthy obese phenotype), and those within ±1 SD as “VAT BMI-Expected.” The same thresholds were applied on the VAT-HbA1c axis, producing “VAT Above HbA1c-Expected” (visceral adiposity exceeding HbA1c’s prediction), “VAT Below HbA1c-Expected” (elevated glycemic markers without proportionate visceral adiposity), and “VAT HbA1c-Expected” groups. Cross-discordance was assessed by classifying participants with both VAT-BMI and VAT-HbA1c assignments into their joint group membership. We also examined how many discordant individuals were below the clinical thresholds considered as healthy - BMI < 25 kg/m^2^ (normal weight) and HbA1c < 5.7% (normoglycemic). Between-group differences across 17 clinical and CGM features were assessed by Kruskal-Wallis test with Benjamini-Hochberg FDR correction. As a sensitivity analysis, we repeated the analyses while employing a threshold of ±2 SD.

To study whether metabolomics can predict IR, we used GBR^36^ with 5-fold cross-validation to predict each IR surrogate from metabolomics features. Missing values were imputed with column median within each cross-validation fold. SHAP feature importance^37^ was computed on models trained on the full dataset. For discordance discrimination, metabolite levels were compared across residual-based discordance groups (e.g., VAT Above BMI-Expected / VAT BMI-Expected / VAT Below BMI-Expected) using Kruskal-Wallis tests with Benjamini-Hochberg FDR correction.

To test whether CGM parameters discriminate the residual-based discordance groups defined above, we computed three complementary analyses on the 6,777 VAT-BMI and 1,981 VAT-HbA1c participants with both discordance-group assignment and CGM coverage. First, each of the 29 IGLU CGM metrics was tested across the three groups on each axis by Kruskal-Wallis with Benjamini-Hochberg FDR correction, and we report per-metric Cohen’s d versus the expected reference group using pooled standard deviation. Second, we computed per-metric AUC for each of the four discordant-vs-expected binary contrasts (VAT Above BMI-Expected, VAT Below BMI-Expected, VAT Above HbA1c-Expected, VAT Below HbA1c-Expected) with 1,000-iteration stratified bootstrap 95% CIs. Third, to quantify CGM information content beyond mean glycemia - particularly relevant on the VAT-HbA1c axis where CGM mean glucose, GMI, and eA1c are HbA1c-correlated, we fit two logistic-regression models per contrast with 5-fold stratified cross-validation after median imputation and z-scoring: Model A used only variability-based CGM metrics (MAGE, CV, MODD, CONGA, GVP, LBGI, HBGI, ADRR), and Model B added glucose mean. Bootstrap 95% CIs (1,000 iterations) were computed on the out-of-fold probability predictions.

To validate associations between CGM metrics and IR surrogates we analyzed Spearman correlations in the 1:1 caliper-matched HPP subcohort (N = 135) and the matched PPT subcohort (N = 135). Replication was assessed by Pearson correlation between the 261 matched effect-size pairs present in both cohorts.To validate IR surrogates against complementary reference measures, we computed Spearman correlations between available surrogates and HOMA-IR within the PPT cohort (N = 165, where fasting insulin was available) and between the same surrogates and DEXA VAT mass within the matched HPP subcohort (N = 66-88, depending on surrogate availability within the 135-participant matched subset).

All correlations presented in the paper are Spearman rank unless otherwise specified. Bootstrap confidence intervals used 1,000 iterations with replacement. Predictive model performance is reported as cross-validated R^2^ and Spearman rho between predicted and observed values. Between-group comparisons used Kruskal-Wallis tests; multiple comparison correction used the Benjamini-Hochberg procedure throughout. All analyses were performed in Python 3.12 using scipy 1.11, scikit-learn 1.3, and SHAP 0.43.

### Ethics approval

Written informed consent was obtained from all participants upon arrival at the research site. Participant identifying information was removed before any computational analysis was performed. The HPP cohort study was conducted in accordance with the Declaration of Helsinki and received approval from the Institutional Review Board (IRB) of the Weizmann Institute of Science (approval number 2392-4)

## Results

### Normative Distributions and Sex- and Age-Specific Patterns of IR Surrogates in Non-Diabetic Adults

Following the exclusion process (see Methods, Supplementary Figure S1), 10,114 non-diabetic adults were included in the analysis (53.3% female, mean age 51.4±7.8 years, mean BMI 25.8±4 kg/m^2^) (Table 2).

**Table 2.**
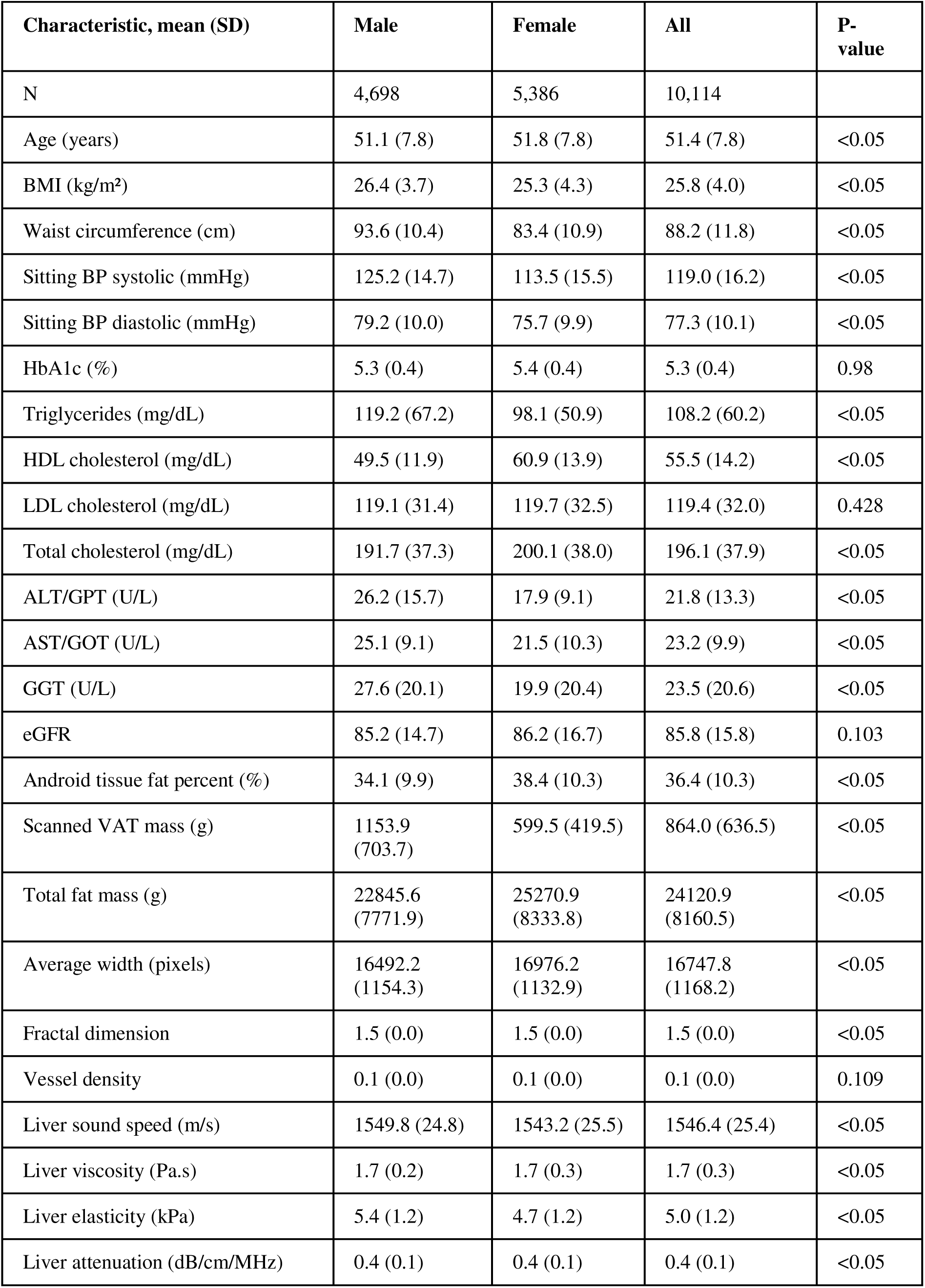
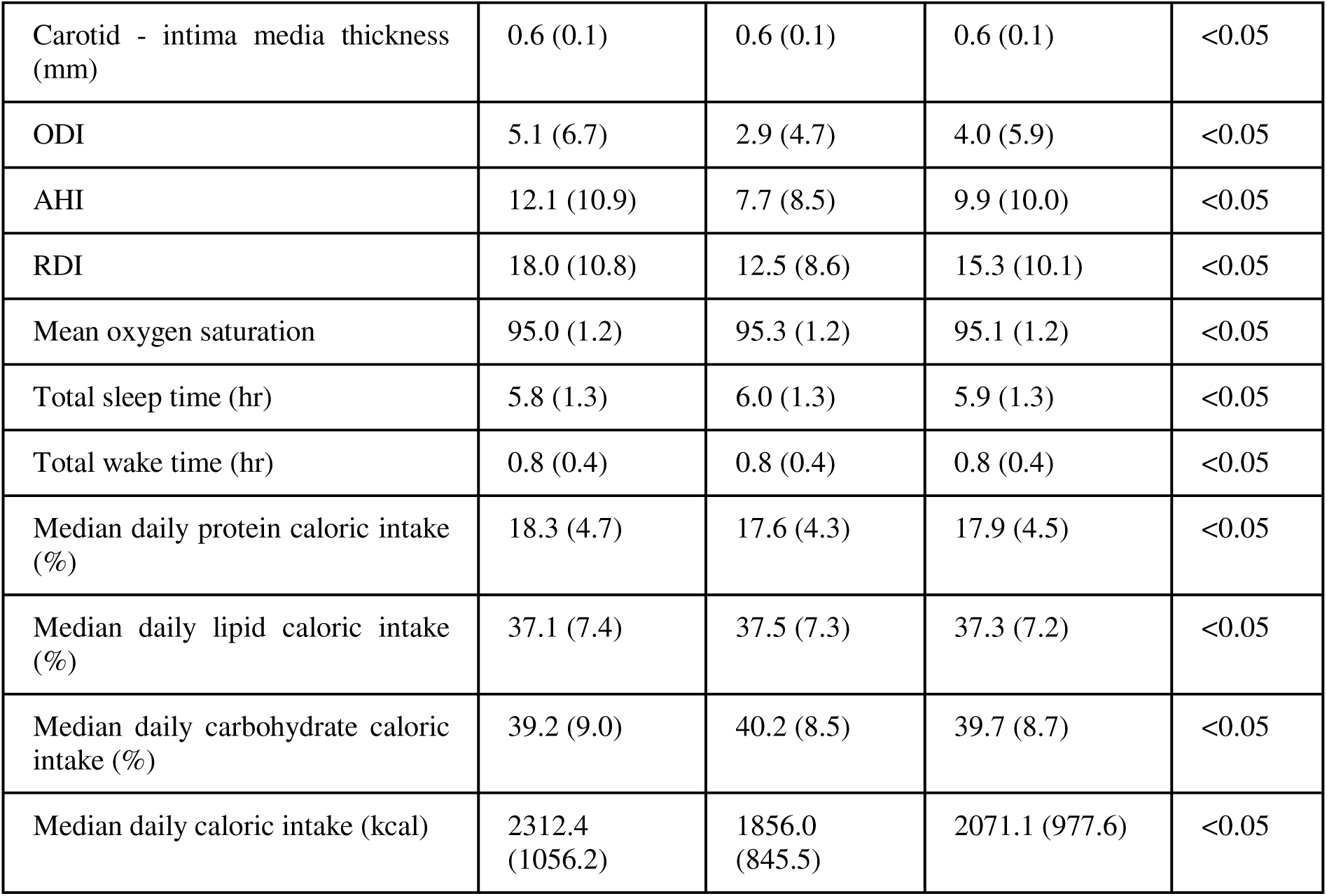
Baseline characteristics of the study population. Sex-stratified baseline characteristics of the 10,114 non-diabetic adults in the final analytic cohort. Values presented as mean (SD) or N (%). P-values for sex differences by Kruskal-Wallis test. Abbreviations: AHI — apnea-hypopnea index; ALT/GPT — alanine aminotransferase/glutamic-pyruvic transaminase; AST/GOT — aspartate aminotransferase/glutamic-oxaloacetic transaminase; BMI — body mass index; BP — blood pressure; eGFR — estimated glomerular filtration rate; GGT — gamma-glutamyl transferase; HbA1c — glycated hemoglobin; HDL — high-density lipoprotein; LDL — low-density lipoprotein; ODI — oxygen desaturation index; RDI — respiratory disturbance index; SD — standard deviation; VAT — visceral adipose tissue.

We selected IR surrogate measures available in the HPP cohort ^16^ validated to varying degrees against the euglycemic hyperinsulinemic clamp (EHC) by existing medical literature (See Methods, Table 1). HOMA-IR, as well as other measures based on fasting insulin levels ^23^, were unavailable due to absent fasting insulin data in the HPP cohort. We therefore focused on non-insulin based measures, emulating real life clinical settings in which insulin levels are not routinely measured, and validated our findings on an independent validation cohort in which fasting insulin levels were available ^20^.

IR surrogates analyzed included anthropometric indices - BMI, Waist Circumference (WC), and the Lipid Accumulation Product (LAP), representing simple, clinically accessible measures of adiposity; Biochemical and metabolic composites included the Triglyceride-Glucose Index (TyG), TyG-BMI, the Metabolic Score for Insulin Resistance (METS-IR), the Triglyceride-to-HDL Cholesterol Ratio (TG/HDL), and Fasting Plasma Glucose (FPG), each derived from routine laboratory values; and Imaging-based measures included DEXA-derived VAT, the Android-to-Gynoid Fat Ratio (A/G Ratio), and Liver Attenuation assessed by ultrasound-controlled attenuation parameter (CAP), reflecting fat distribution and ectopic lipid deposition as physiological correlates of IR. Together, these measures represent a broad spectrum, from universally available clinical tests to imaging assessments, allowing a comprehensive comparison of their relative utility in capturing insulin resistance across the study population.

The distributions of the selected surrogate measures are shown in Figure 1A (distribution of BMI, waist circumference, and fasting glucose are shown in Figure S2). All surrogates showed approximately unimodal distributions with right skew for adiposity-based and lipid-derived measures, consistent with the known positive skew of triglyceride distributions in general population samples ^38^. The cohort displayed substantial heterogeneity across all IR surrogate measures despite uniform exclusion of diabetes, underscoring that non-diabetic status does not imply metabolic homogeneity.

**Figure 1.**
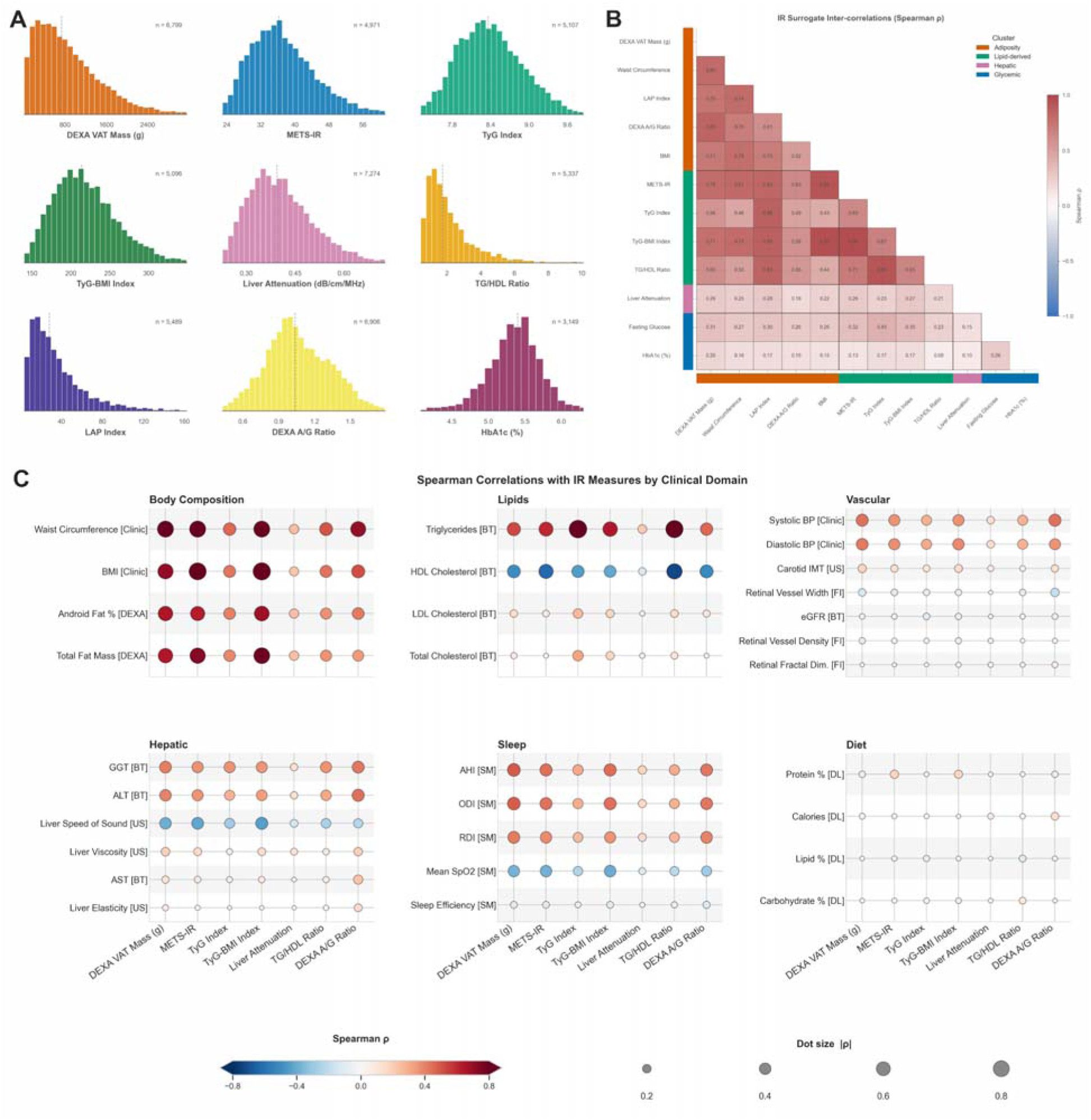
IR measures landscape. (A) Distributions of 9 IR surrogate measures (BMI, fasting glucose, and waist circumference shown separately in Supplementary Figure S2). (B) Spearman inter-correlation matrix of IR surrogate measures, ordered by cluster (adiposity, lipid-derived, hepatic, glycemic; colored side bars and legend) and by clamp-rank within cluster. (C) Dot-matrix of Spearman correlations between clinical variables and IR surrogates across 6 clinical domains; dot size encodes Spearman rho, color encodes signed rho.

Men had approximately two-fold higher VAT mass (1,153.9 ± 703.7 g vs. 599.5 ± 419.5 g, p < 0.001), higher waist circumference, triglycerides, METS-IR, and TyG index, while women had higher HDL cholesterol and total body fat percentage (all p < 0.001; Table 1). These differences were not uniform across surrogate categories: lipid-derived surrogates (TG/HDL ratio, LAP index) showed larger sex differences than glycemic surrogates (HbA1c, fasting glucose), confirming that different surrogate domains are differentially sensitive to sex-related metabolic variation. Reference values for the different IR surrogates are presented in Table 1.

Age and sex related progression (Supplementary Figure S3) showed a consistent pattern of increasing IR burden across adiposity-based and lipid-derived surrogates with advancing age, with steeper trajectories in men for VAT-based measures and more attenuated trajectories in women, particularly in the post-menopausal age range represented in this cohort. Glycemic surrogates showed more modest age-related increases.

### IR Surrogate Measures Capture Distinct Metabolic Facets and Are Moderately Inter-Correlated

The IR surrogate measures exhibited moderate inter-correlations with a clear cluster structure across four categories (indicated by the colored side bars): adiposity-based (DEXA VAT mass, waist circumference, LAP index, DEXA A/G ratio, BMI), lipid-derived composites (METS-IR, TyG index, TyG-BMI, TG/HDL ratio), hepatic (liver attenuation), and glycemic (fasting glucose, HbA1c) (Figure 1B). Within-cluster correlations were strongest for adiposity-based measures (pairwise Spearman ρ: 0.52–0.83) and lipid-derived composites (ρ: 0.63–0.96). Several cross-cluster associations were also high, largely because lipid-derived composites explicitly incorporate adiposity components: METS-IR with BMI (ρ = 0.92), TyG-BMI with BMI (ρ = 0.95), and DEXA VAT mass with METS-IR (ρ = 0.79). In contrast, hepatic and glycemic surrogates showed weak correlations with other measures (|ρ| ≤ 0.40), and the two glycemic markers were only modestly correlated with each other (ρ = 0.26). Overall, this hierarchical structure suggests that while these surrogates capture related aspects of metabolism, they reflect partially distinct facets, and no single measure fully represents the spectrum of IR pathophysiology.

Clinical domain correlations across six organ systems confirmed this hierarchical structure (Figure 1C). Body composition markers showed the strongest associations with IR surrogates across domains. Hepatic markers (GGT, ALT) correlated preferentially with adiposity-based surrogates, consistent with the role of visceral adipose tissue in driving portal free fatty acid flux and hepatic steatosis ^39^. Vascular markers (blood pressure, carotid IMT) showed moderate associations across surrogate categories. Sleep-disordered breathing indices showed strong associations with visceral adiposity, consistent with the bidirectional relationship between obstructive sleep apnea and metabolic syndrome ^40^. Dietary parameters showed the weakest correlations with IR surrogates. Overall, DEXA-derived VAT mass emerged as the most informative cross-domain surrogate in this cohort, showing significant associations with body composition, hepatic, vascular, lipid, and sleep-related markers. Abate et al.^25^ demonstrated, using CT-measured visceral fat and the Euglycemic–Hyperinsulinemic Clamp, that intra-abdominal fat was the strongest adiposity-based predictor of insulin-mediated glucose disposal (r −0.76 with M-value). Additionally, DEXA-derived VAT has been shown to correlate with clamp-measured insulin sensitivity in both children and adults ^26,27^.

Accordingly, we selected VAT mass as the primary surrogate of insulin resistance for the subsequent analyses presented in this study.

### VAT Mass Can Be Accurately Estimated from Routine Clinical Data

Given that DEXA imaging is not universally available, we also examined whether VAT mass could be accurately estimated from routinely available clinical measurements. To facilitate clinical implementation, we fit both gradient-boosting and interpretable linear models using 5-fold cross-validation (N = 6,799 participants with both DEXA and clinical data). An anthropometric model which includes sex, age, blood pressure, BMI, weight, and height achieved cross-validated R^2^ - 0.659 (rho = 0.802). The addition of waist circumference, hip circumference, and waist-to-hip ratio further improved performance to R^2^ -0.729 (rho = 0.840). In the circumference model, waist circumference emerged as the dominant predictor, confirming that central adiposity distribution, rather than total body mass, drives VAT estimation accuracy.

### Glycemic Variability Metrics Outperform Mean Glucose in Capturing Visceral Adiposity

The association between IR and CGM-derived parameters in individuals without diabetes is scarcely studied. Examining the relationship between 29 CGM-derived metrics and IR surrogates (Figure 2A shows a representative 10-metric subset; full set in Supplementary Figure S4), we identified modest yet statistically significant correlations between them (up to r = 0.3). A gradient boosting model using 29 CGM features, sex, and age achieved cross-validated R^2^ = 0.276 (Spearman rho = 0.524) for predicting DEXA VAT mass (N = 6,799; Supplementary Figure S5). Because ∼2-fold sex differences in VAT alone carry a substantial share of this predictive weight, we tested a model using only the 29 CGM features: this yielded cross-validated R^2^ = 0.078 (Spearman rho = 0.291), indicating that CGM alone encodes only a modest signal for VAT prediction. SHAP feature importance analysis on the full CGM + sex + age model revealed that glucose variability metrics dominated over glucose level metrics (Figure 2B). Sex was the dominant predictor overall (22.5% of total SHAP importance), consistent with the approximately two-fold sex difference in VAT mass. Among CGM features, MAG (7.3%), SD of rate-of-change (6.9%), and MODD (6.6%) were the top contributors, collectively accounting for 20.8% of total SHAP importance, while mean glucose and GMI contributed minimally. Subgroup analyses confirmed that CGM-IR associations strengthened with metabolic burden: correlations were larger in obese individuals (BMI >= 30, N = 1,435) and those with prediabetic HbA1c (HbA1c 5.7-6.5%, N = 587), suggesting that CGM-derived metrics become increasingly informative as underlying metabolic burden increases (Supplementary Figures S6).

**Figure 2.**
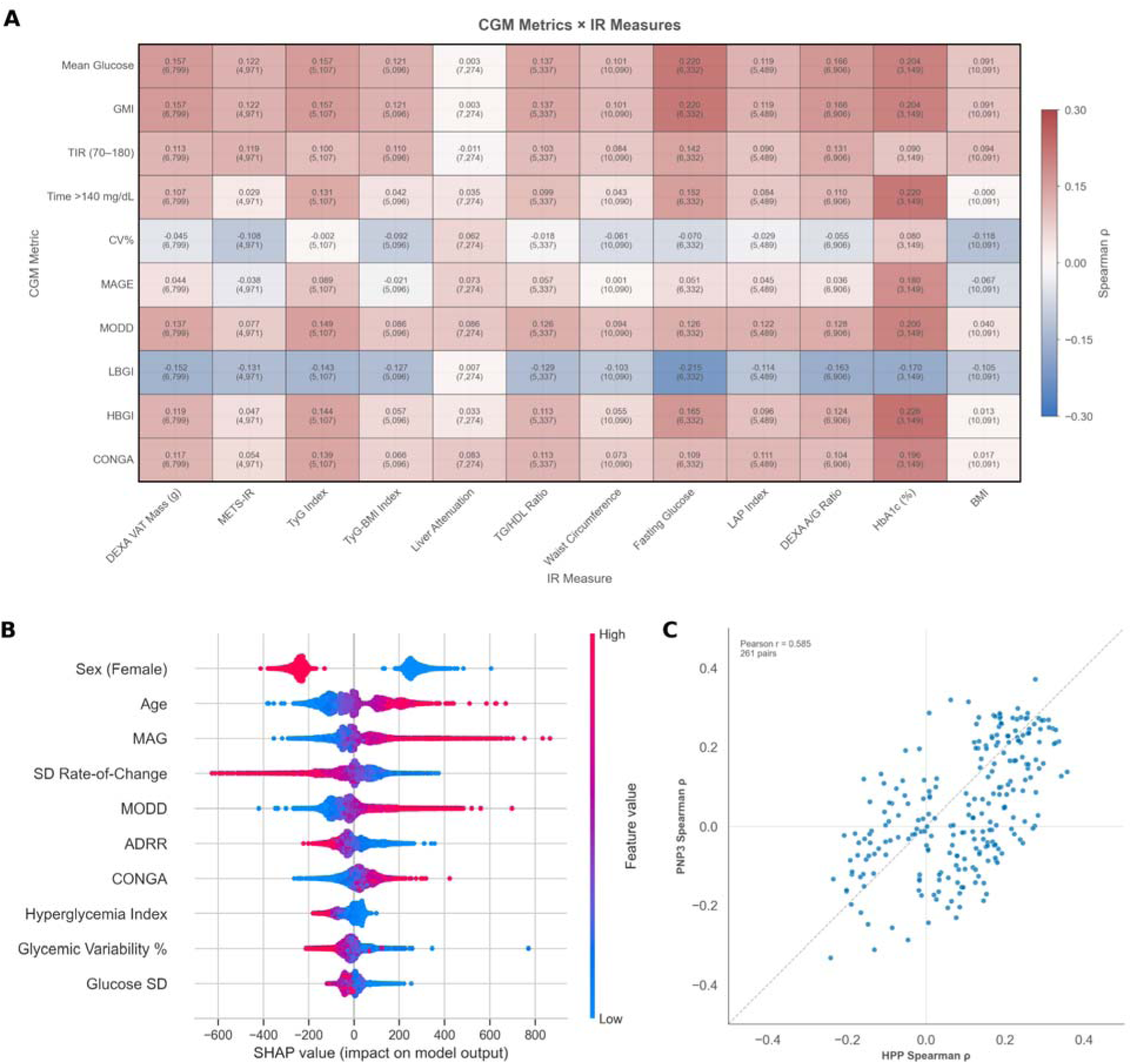
CGM associations with IR measures and CGM-based prediction of visceral adiposity. (A) Heatma of Spearman correlations between a representative 10-metric subset of the CGM-derived features and core IR measures (full 29-metric heatmap in Supplementary Figure S4). Cell annotations show rho and sample size. (B) SHAP plot showing the 10 most impactful features for VAT mass prediction. Each point represents one participant; color indicates feature value. The corresponding predicted-vs-actual scatter (N = 6,799; R^2^ = 0.276, Spearman rho = 0.524) is shown in Supplementary Figure S5. (C) Replication of CGM-IR effect sizes between HPP and PPT cohorts. Abbreviations: ADRR — average daily risk range; AHI — apnea-hypopnea index; A/G — android-to-gynoid; BMI — body mass index; CGM — continuous glucose monitoring; CONGA — continuous overall net glycemic action; CV% — coefficient of variation; DEXA — dual-energy X-ray absorptiometry; eGFR — estimate glomerular filtration rate; GMI — glucose management indicator; HbA1c — glycated hemoglobin; HBGI — high blood glucose index; HDL — high-density lipoprotein; HPP — Human Phenotype Project; IR — insulin resistance; LAP — lipid accumulation product; LBGI — low blood glucose index; MAG — mean absolute glucose change; MAGE — mean amplitude of glycemic excursions; METS-IR — metabolic score for insulin resistance; MODD — mean of daily differences; PPT — Personalized Postprandial Glucose Response-Targeting Diet; SD — standar deviation; SHAP — Shapley additive explanations; TG — triglycerides; TIR — time in range; TyG — triglyceride-glucose index; VAT — visceral adipose tissue.

### Residual-Based Discordance Analysis Reveals Metabolically At-Risk Individuals Missed by BMI and HbA1c Screening

We next examined whether conventional clinical classification by BMI or HbA1c fails to identify individuals with elevated visceral adiposity and adverse metabolic profiles. Using a residual-based approach (Figure 3A,B, see Methods), individuals were classified by their standardized residual: those exceeding +1 SD were designated as “VAT Above BMI-Expected” (visceral adiposity higher than their BMI predicts; analogously “VAT Above HbA1c-Expected” for the HbA1c axis), those below -1 SD as “VAT Below BMI-Expected” / “VAT Below HbA1c-Expected,” and those within ± 1SD as “VAT BMI-Expected” / “VAT HbA1c-Expected.” Among 6,777 participants with both DEXA and BMI data, 987 (14.6%) were classified as VAT Above BMI-Expected, 4,818 (71.1%) as VAT BMI-Expected, and 972 (14.3%) as VAT Below BMI-Expected.

**Figure 3.**
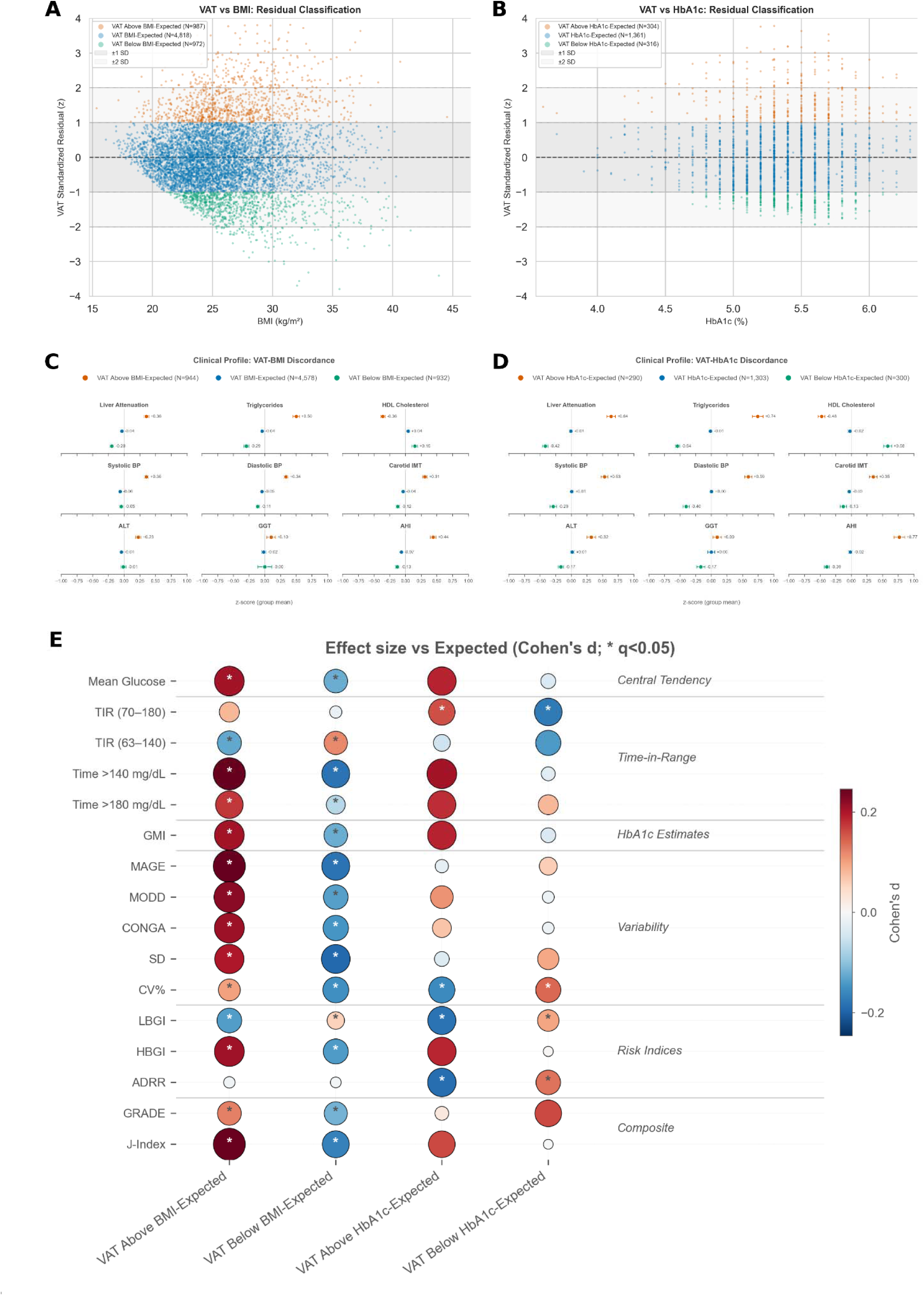
Residual-based discordance between VAT and conventional markers. (A, B) Standardized-residual scatter plots (y-axis = VAT residual z-score from sex-specific regression on BMI [A] or HbA1c [B]); horizontal line at 0 is the regression fit, shaded bands mark the ±1 SD and ±2 SD classification thresholds, and points are color-coded by discordance group. (C, D) Small-multiples forest of z-scored group means (± SE) for directly measured clinical parameters (liver attenuation, triglycerides, HDL, SBP, DBP, carotid IMT, ALT, GGT, AHI), ordered by organ system (Adipose/Lipid, Vascular, Hepatic, Sleep) on a shared z-score x-axis across panels C and D. (E) CGM effect-size matrix: dot-matrix of Cohen’s d for a curated subset of 16 selected IGLU CGM metrics (variability, risk, time-in-range, and composite categories) across four discordant-vs-expected contrasts; dot area encodes Cohen’s d|, colour encodes sign, star denotes q < 0.05). Top discriminating NMR metabolites between discordant subphenotypes are shown in Figure 4C. All between-group comparisons significant at FDR < 0.05. Abbreviations: ADRR — average daily risk range; AHI — apnea-hypopnea index; ALT — alanine aminotransferase; BMI — body mass index; BP — blood pressure; CONGA — continuous overall net glycemic action; CV% — coefficient of variation; GGT — gamma-glutamyl transferase; GMI — glucose management indicator; GRADE — glycemia risk assessment diabetes equation; HbA1c — glycated hemoglobin; HBGI — high blood glucose index; HDL — high-density lipoprotein; IMT — intima-media thickness; LBGI — low blood glucose index; MAGE — mean amplitude of glycemic excursions; MODD — mean of daily differences; SD — standard deviation; TIR — time in range; VAT — visceral adipose tissue.

The VAT Above BMI-Expected group, which includes individuals whose visceral adiposity substantially exceeds what their BMI would predict, capturing the TOFI (“thin outside, fat inside”) phenotype, showed significantly worse metabolic profiles than the VAT BMI-Expected group across all clinical features examined (Figure 3B, Kruskal-Wallis p < 0.05, FDR-corrected), and had elevated IR surrogates- METS-IR, TyG index, TG/HDL ratio, LAP index, and systolic blood pressure (Supplementary Figure S7A). Conversely, the VAT Below BMI-Expected group, which includes individuals with lower visceral adiposity than their BMI predicts, showed a more benign metabolic profile than their BMI would suggest.

Among 1,981 participants with both DEXA and HbA1c data, 304 (15.3%) were classified as VAT Above HbA1c-Expected (VAT higher than HbA1c predicts), 1,361 (68.7%) as VAT HbA1c-Expected, and 316 (16.0%) as VAT Below HbA1c-Expected (elevated HbA1c without proportionate visceral adiposity). Notably, HbA1c is only weakly correlated with VAT in this cohort (Spearman rho = 0.204). The VAT Above HbA1c-Expected group showed elevated adiposity and lipid markers, with significant between-group differences across all examined features (Figure 3D, Supplementary Figure S7B).

Cross-discordance analysis classified on both axes revealed substantial coupling between the two forms of discordance: 168 of 294 (57%) VAT Above BMI-Expected individuals were also VAT Above HbA1c-Expected, and 168 of 303 (55%) VAT Above HbA1c-Expected individuals were also VAT Above BMI-Expected, consistent with a shared underlying visceral-IR phenotype. While the two residual axes provide partially overlapping complementary lenses rather than orthogonal ones, their disagreement on 43-45% of dual-classified cases means that using either marker alone still fails to flag a substantial fraction of the at-risk population.

We next examined the subset of discordant individuals who fall within conventional normal ranges on both BMI (< 25 kg/m^2^) and HbA1c (< 5.7%) and would therefore pass both screening thresholds despite elevated visceral adiposity. Among the 987 VAT Above BMI-Expected individuals, 337 (34.1%) had BMI < 25 and 207 (21.0%) had HbA1c < 5.7%; while 80 individuals (1.2% of 6,777 VAT-BMI classifiable participants) met both criteria simultaneously (mean VAT: 1,155 g, mean BMI: 23.3, mean HbA1c: 5.3%). Among the 304 VAT Above HbA1c-Expected individuals, only 9 individuals (0.4% of 1,981) were screening-invisible by both criteria simultaneously. These individuals represent a population in whom visceral IR has advanced sufficiently to produce adverse metabolic consequences across multiple organ systems, yet who remain below the BMI and HbA1c threshold that would trigger clinical concern.

As a further sensitivity analysis, we repeated the residual-based approach using a stricter ±2 SD threshold. On the VAT-BMI axis, this identified 233 (3.4%) VAT Above BMI-Expected and 97 (1.4%) VAT Below BMI-Expected individuals. On the VAT-HbA1c axis, 83 (4.2%) were classified as VAT Above HbA1c-Expected. Metabolic profile comparisons remained significant at FDR < 0.05 for these extreme groups (Supplementary Figure S8), confirming that findings are robust to the choice of threshold.

### CGM Variability Discriminates Discordant VAT Phenotypes and Carries Signal Beyond Mean Glycemia

Having defined discordant VAT subphenotypes on the BMI and HbA1c axes (Figure 3A–D), we tested whether CGM metrics can discriminate between these groups. Across 29 CGM-derived metrics, 26 (90%) showed FDR-significant differences across the three VAT-BMI groups (n = 6,777), and 10 (34%) across the three VAT-HbA1c groups (Figure 3E). On both axes, the largest between-group effect sizes were concentrated among variability metrics, whereas mean glucose and its derivatives (GMI, eA1c) were among the least informative, particularly on the HbA1c axis, where these metrics are structurally constrained by the variable used to define the groups.

To formally test whether variability carries discriminative information beyond mean glucose, we compared two logistic regression models per contrast (5-fold cross-validation, bootstrap 95% CI; Supplementary Figure S9): Model A included eight variability metrics (MAGE, CV, MODD, CONGA, GVP, LBGI, HBGI, ADRR); Model B added mean glucose. For the most clinically relevant contrast, VAT Above HbA1c-Expected versus VAT HbA1c-Expected, Model A achieved AUC = 0.632 (95% CI 0.598–0.666; n = 304 vs. 1,361), while Model B yielded AUC = 0.636 (ΔAUC = +0.003). This negligible increment was consistent across all four contrasts (VAT-BMI axis: ΔAUC = −0.002 and +0; Supplementary Figure S9B). At the single-metric level, variability measures (SD of rate-of-change, GVP, MODD) consistently outperformed mean glucose, with top AUCs of 0.55–0.56 (Supplementary Figure S9A). These effects demonstrate that two-week CGM captures information about visceral adiposity beyond what HbA1c provides, with the signal residing in glycemic dynamics rather than levels.

### Metabolomic Correlates of Insulin Resistance and Their Distribution Across Discordant VAT Subphenotypes

We next analyzed whether NMR metabolomics data by Nightingale Health platform are associated with IR heterogeneity metrics and can capture discordant subphenotypes identified above (see Methods). As expected from the fact that lipoprotein subfractions are included in the NMR panel, Lipid-derived and adiposity-based IR surrogates were predicted with high accuracy: TyG index (R^2^=0.580), TG/HDL ratio (R^2^ =0.570), METS-IR (R^2^=0.509), A/G ratio (R^2^=0.509), and DEXA VAT mass (R^2^=0.475). Additional targets included waist circumference (R^2^=0.396) and liver attenuation (R^2^=0.043). SHAP feature importance analysis identified VLDL and HDL particle subfractions as the dominant metabolite predictors across lipid-derived and adiposity-based targets (Figure 4B), consistent with the established roles of atherogenic dyslipidemia and impaired reverse cholesterol transport in IR pathophysiology ^41^. Amino acids and glycoprotein acetyls contributed to adiposity-based predictions, potentially reflecting IR-associated alterations in protein catabolism and low-grade inflammation ^42^.

**Figure 4.**
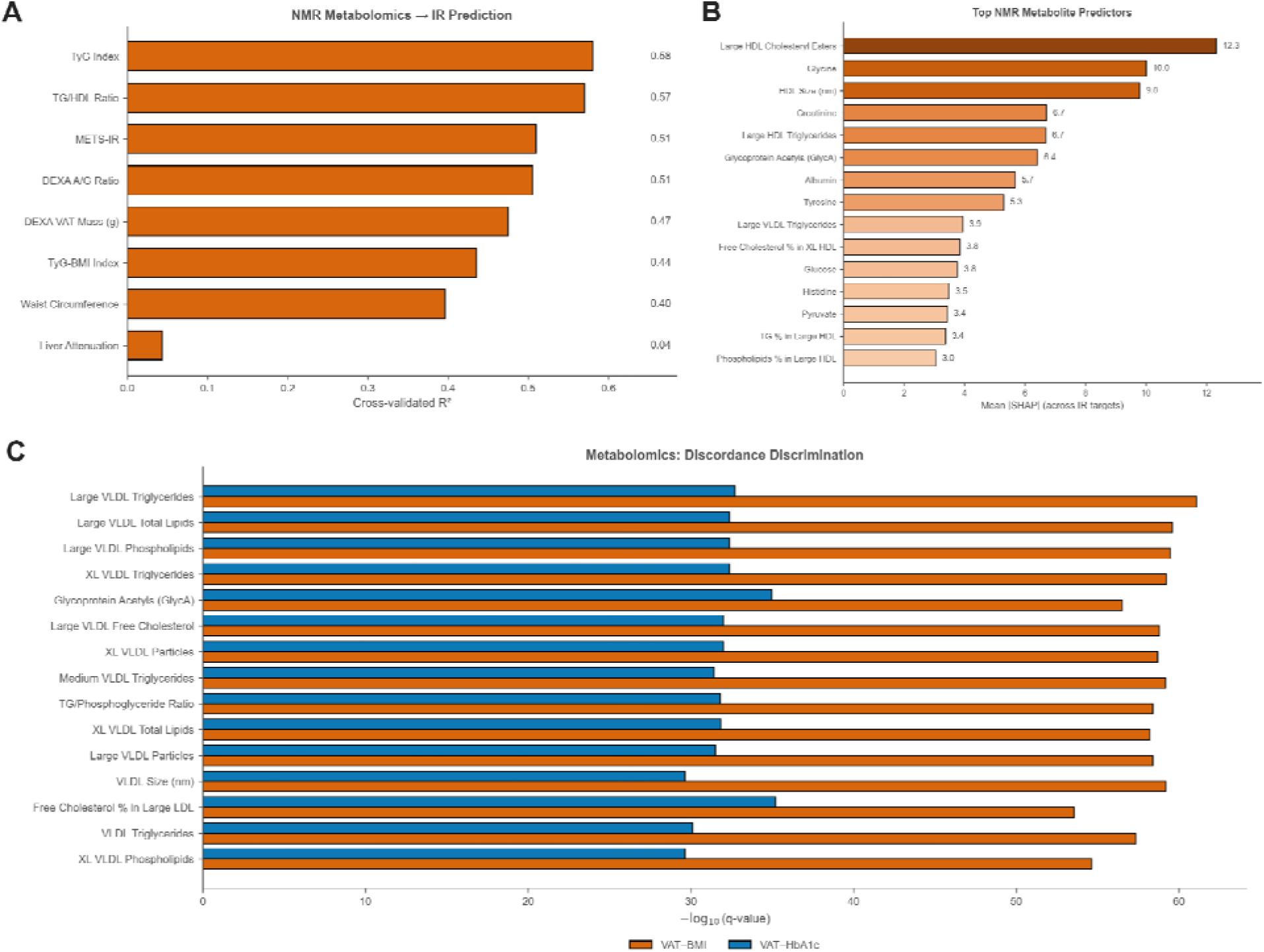
NMR metabolomics: IR prediction, feature importance, and discordance discrimination. (A) Cross-validated R^2^ for gradient boosting models predicting each IR surrogate from ∼250 NMR metabolomics features. (B) Top 15 NMR metabolite predictors across IR targets, ranked by mean |SHAP| value. (C) Top NMR metabolites discriminating discordant VAT subphenotypes on both VAT-BMI (orange) and VAT-HbA1c (blue) axes, ranked b-log10(q-value) from Kruskal-Wallis tests with Benjamini-Hochberg FDR correction. Lipoprotein subfractions (VLDL and HDL triglyceride and phospholipid content) dominate discrimination on both axes.

NMR metabolites discriminated between residual-based discordance groups on both the VAT-BMI and VAT-HbA1c axes (FDR < 0.05; Figure 4C). The top-ranked discriminators on both axes were large VLDL and large/very-large HDL triglyceride and phospholipid subfractions (Figure 4C), consistent with the VLDL-overproduction / HDL-remodeling axis driven by portal free-fatty-acid flux in visceral IR ^43^. VAT Above BMI-Expected individuals differed metabolomically from VAT BMI-Expected individuals in these lipoprotein subfraction profiles and in glycoprotein acetyls, despite similar BMI classification. VAT Above HbA1c-Expected individuals similarly showed a distinct metabolomic signature relative to the VAT HbA1c-Expected group despite identical glycemic classification.

### Validation of IR Surrogates in an Independent Cohort with Fasting Insulin Data

To validate the IR surrogates used throughout this study, we leveraged the PPT cohort (n = 165 participants with available HOMA-IR, see Methods). Among individual surrogates, METS-IR showed th strongest correlation with HOMA-IR (Spearman ρ = 0.574), followed by LAP index (ρ = 0.572), TyG-BMI (ρ = 0.566), waist circumference (ρ = 0.507), and BMI (ρ = 0.501). Lipid-glucose indices showed intermediate correlations (TG/HDL ratio ρ = 0.446; TyG index ρ = 0.440). Although individual surrogate correlations with HOMA-IR were modest, a multivariable linear regression combining all available surrogates yielded R² = 0.247 (ρ = 0.603, n = 165), substantially exceeding any single surrogate. Thi confirms that each measure captures partially non-overlapping IR information, consistent with th moderate inter-surrogate correlations observed in the HPP cohort.

We further assessed cross-cohort replicability of CGM–IR associations by comparing 261 matched correlation pairs (29 CGM metrics × 9 IR surrogates available in both cohorts). Effect sizes replicated with Pearson r = 0.585 between HPP and PPT estimates, with 172 of 261 pairs (66%) showing directionally consistent associations (Figure 2C). Mechanistically relevant variability associations, including MODD × TyG index and MODD × HbA1c, replicated in both direction and relative magnitude across cohorts, confirming that CGM-derived glycemic variability captures metabolically meaningful signals that generalize beyond a single population.

## Discussion

This study provides a comprehensive, multi-modal characterization of insulin resistance heterogeneity in over 10,000 non-diabetic adults, integrating validated IR surrogates, DEXA body composition, 14-day continuous glucose monitoring, and NMR metabolomics. Our findings yield four principal insights: (i) IR surrogates capture related but distinct metabolic facets, with no single measure approximating the full spectrum of IR pathophysiology; (ii) visceral adipose tissue can be accurately estimated from routine anthropometry, and CGM-derived glycemic variability outperforms mean glucose in capturing visceral adiposity; (iii) a clinically important subset of individuals with normal BMI and HbA1c harbor elevated visceral fat and multi-organ metabolic adversity; and (iv) NMR lipoprotein subfractions and CGM variability metrics discriminate these discordant phenotypes, offering complementary inputs for early metabolic risk detection.

Insulin resistance exists along a continuous spectrum, spanning hepatic, skeletal muscle, and adipose tissue compartments that can be independently and variably impaired ^44^. Our observation that IR surrogates form four partially independent clusters- adiposity-based, lipid-derived, hepatic, and glycemic, with moderate cross-cluster correlations aligns with this framework. Within-cluster correlations were strongest among adiposity-based measures (ρ 0.52–0.83) and among lipid-derived composites (ρ 0.63–0.96). Several cross-cluster pairs were also highly correlated because the lipid-derived composites explicitly embed an adiposity term (e.g., METS-IR and BMI, ρ = 0.92; TyG-BMI and BMI, ρ = 0.95). Hepatic and glycemic surrogates were only weakly associated with the rest of the panel (|ρ| ≤ 0.40). This hierarchical structure mirrors the known physiological partitioning of IR into distinct tissue compartments and implies that reliance on any single surrogate will systematically underestimate IR burden in a fraction of the population. The validation analysis in the PPT cohort reinforced this conclusion: while individual surrogate correlations with HOMA-IR were modest (maximum ρ = 0.574 for METS-IR), combining all surrogates yielded R² = 0.247, substantially exceeding any single measure. This additive variance confirms that each surrogate captures partially non-redundant metabolic information.

Our sex-specific reference values for these surrogates, derived from a large non-diabetic population, address a gap in the literature. While individual surrogate measures have published normative data, to the best of our knowledge, comprehensive sex- and age-stratified reference ranges across the full panel of non-insulin-based IR measures have not previously been established within a single deeply phenotyped cohort. These values may serve as benchmarks for clinical interpretation and future research.

Among the surrogates examined, DEXA-derived VAT mass emerged as the most informative cross-domain correlate, showing significant associations with body composition, hepatic, vascular, lipid, and sleep-related markers. This is consistent with the established role of visceral adiposity as the metabolic driver linking portal free fatty acid flux, hepatic steatosis, atherogenic dyslipidemia, and systemic inflammation ^10^ ^45^. The finding that VAT can be predicted from routine anthropometric (R² = 0.659 from sex, age, blood pressure, BMI, weight, and height; R² = 0.729 with the addition of circumference measures) is aligned with previous literature ^46^ and has practical implications: it suggests that clinically meaningful VAT estimation does not require DEXA and could be integrated into routine health assessments. Waist circumference was the dominant predictor in the circumference model, consistent with prior evidence that central adiposity distribution, rather than total body mass, drives cardiometabolic risk ^47^.

A major strength of this study is the availability of CGM data. We show a consistent hierarchy in which CGM-derived glycemic variability metrics (MAG, SD of rate-of-change, MODD) outperformed mean glucose and its derivatives (GMI, eA1c) in capturing visceral adiposity and discriminating discordant VAT phenotypes. This pattern was observed across multiple analytical frameworks: in VAT prediction models (SHAP importance), in cross-sectional correlations with IR surrogates, in between-group effect sizes across discordance groups, and in the logistic regression analysis where adding mean glucose to a variability-only model yielded negligible improvement.

This finding extends prior work showing associations between glycemic variability and cardiometabolic markers in non-diabetic individuals. A systematic review and meta-analysis of 71 studies found that while glycemic variability was inversely associated with beta-cell function, its relationship with insulin sensitivity and adiposity remained inconclusive ^48^. Our study, powered by 6,799 participants with concurrent DEXA and CGM, resolves this ambiguity by demonstrating that CGM variability carries statistically meaningful, replicable information about visceral adiposity beyond what HbA1c provides, though effect sizes are modest (AUC 0.55–0.63). Importantly, CGM-IR associations strengthened with increasing metabolic burden (obesity, prediabetic HbA1c), suggesting that CGM becomes more informative as underlying metabolic dysfunction progresses, consistent with the concept that glycemic variability may reflect an earlier perturbation of insulin-mediated glucose homeostasis before mean glucose rises ^49^.

The mechanistic interpretation of why variability dominates over mean glucose deserves consideration. In the pre-diabetic state, compensatory hyperinsulinemia maintains near-normal mean glucose while glucose excursion dynamics, reflecting the interplay of hepatic glucose output and peripheral uptake kinetics become dysregulated. Variability metrics may therefore capture this dynamic dysregulation that remains invisible to static measures like HbA1c or mean glucose ^50^.

The most clinically consequential finding is the identification of a subpopulation whose visceral adiposity substantially exceeds what their BMI or HbA1c would predict and who harbor adverse metabolic profiles across lipid, hepatic, vascular, and sleep domains, yet remain below both screening thresholds that would trigger clinical attention. These individuals (1.2% of the cohort on the VAT-BMI axis meeting both BMI < 25 and HbA1c < 5.7% criteria simultaneously) represent the clinically occult end of the TOFI phenotype spectrum, first described by Thomas et al. ^12^ and formalized as the metabolically obese normal-weight (MONW) phenotype ^11^.

Our residual-based discordance framework extends prior categorical approaches (e.g., classifying individuals as MONW by fixed BMI and metabolic syndrome criteria) by quantifying the continuous degree of discordance between observed and expected visceral adiposity. The 57% overlap between VAT-BMI and VAT-HbA1c discordance suggests a shared underlying visceral-IR phenotype, while the 43% disagreement indicates that each axis captures a partially distinct dimension of metabolic risk — reinforcing the need for multi-marker screening. A recent Nature Medicine study employing unsupervised clustering to subclassify obesity found that approximately 20% of the general population harbors discordant cardiometabolic profiles that improve risk prediction beyond BMI alone ^51^ consistent with our findings. Our data suggest that complementary markers, particularly waist circumference and lipid-derived indices, substantially improve identification of this at-risk population.

The NMR metabolomics analysis revealed that VLDL and HDL particle subfractions were the dominant discriminators of both IR surrogates and discordant VAT phenotypes. This aligns with the established pathophysiology of atherogenic dyslipidemia in insulin resistance, in which hepatic overproduction of large VLDL1 particles, driven by portal free fatty acid flux, initiates a cascade of lipoprotein remodeling that generates small dense LDL and depletes functional HDL particles ^41^ ^43^). The consistent enrichment of the same VLDL/HDL subfraction discriminators across both discordance axes suggests that a compact targeted lipoprotein panel could identify at-risk individuals currently classified as low-risk by conventional markers, and supports the growing recognition that lipoprotein particle profiles carry metabolic information beyond standard lipid panels ^52^.

Glycoprotein acetyls, a composite NMR-derived marker of systemic low-grade inflammation, contributed to adiposity-based predictions, consistent with the known role of chronic inflammation in linking visceral adiposity to insulin resistance and cardiometabolic disease ^53^. The emergence of amino acids as predictors of adiposity-based IR targets aligns with the established association between branched-chain amino acid catabolism and insulin resistance ^42^, and suggests that NMR metabolomics captures both the lipid and proteomic dimensions of IR heterogeneity.

Taken together, our findings argue for a shift from single-marker to multi-modal metabolic risk assessment. The current clinical paradigm in which BMI and HbA1c serve as primary gatekeepers for metabolic risk may be insufficient to detect a meaningful fraction of non-diabetic adults with visceral IR and multi-organ metabolic adversity. We propose that an extended screening strategy could integrate: (1) anthropometry-based VAT estimation (particularly waist circumference), which is free, universally available, and explained 73% of DEXA VAT variance in our data; (2) composite lipid-glucose indices such as METS-IR or TyG-BMI, which outperform individual markers and can be computed from routine blood work; (3) targeted NMR lipoprotein subfraction profiling, which discriminates discordant phenotypes with high specificity; and (4) CGM-derived glycemic variability metrics, which provide a non-invasive, wearable-based assessment of dynamic glucose regulation that captures metabolic information orthogonal to HbA1c.

Several limitations warrant consideration. First, HOMA-IR was unavailable in the HPP cohort due to absent fasting insulin data, precluding direct comparison of surrogate rankings against this widely used reference. However, validation in the PPT cohort, where HOMA-IR was available, confirmed surrogate rankings and the additive variance structure. Second, the cross-sectional design precludes causal analyses; whether the discordant phenotypes identified here progress to diabetes or cardiovascular disease at differential rates requires prospective follow-up. Third, the cohort is drawn from an Israeli population aged 35–75, and generalizability to other ethnicities and age ranges requires confirmation. Finally CGM discriminative performance for discordant phenotypes, while statistically significant and replicable, yielded modest AUCs (0.55–0.63) insufficient for standalone screening, and should be interpreted as complementary input within a multi-modal framework.

In Summary, this study establishes population-level reference data for non-insulin-based IR surrogates, demonstrates that these surrogates capture complementary facets of a multi-organ condition, and identifies a clinically silent subpopulation with elevated visceral adiposity that escapes detection by conventional BMI and HbA1c screening. CGM-derived glycemic variability and NMR lipoprotein subfractions emerge as promising complementary tools for early metabolic risk stratification, supporting a multi-modal approach to identifying insulin resistance before diagnostic thresholds are crossed. As wearable CGM technology becomes increasingly accessible and NMR metabolomics costs continue to decline, integrating these modalities into preventive medicine frameworks may enable earlier, more precise intervention in the trajectory from metabolic health to type 2 diabetes.

## Declarations

## Acknowledgments

We thank the members of the Segal group for fruitful discussions. S.S. is supported by Dr. Gilbert S. Omenn and Martha A. Darling Weizmann Institute - Schneider Hospital Fund for Clinical Breakthroughs through Scientific Collaborations. E.S. is supported by Dr. Gilbert S. Omenn and Martha A. Darling Weizmann Institute - Schneider Hospital Fund for Clinical Breakthroughs through Scientific Collaborations, Moross Integrated Cancer Center, Swiss Society Institute for Cancer Prevention Research, Seerave Foundation ; and grants funded by the Minerva foundation with funding from the Federal German Ministry for Education and Research and by the European Research Council and the Israel Science Foundation, Israel Ministry of Science. In addition, Louis H. Sackin Research Fellow Chair in Computer Science supports a staff scientist in Prof. Segal’s Lab.

## Ethical considerations

Written informed consent was obtained from all participants at the study site prior to enrollment. Personal identifiers were stripped from the dataset before any computational analysis was performed. The study protocol adheres to the Declaration of Helsinki and was approved by the Weizmann Institute of Science Institutional Review Board (protocol 2392-4).

## Data availability

Due to privacy and ethical considerations set forth by the IRB committee of the study, all other datasets that are part of the Human Phenotype Project (HPP) - such as blood test results and anthropometrics measurements - are accessible to researchers from universities and other research institutions at: https://humanphenotypeproject.org/data-access using the following procedure. Interested bona fide researchers should contact who will then provide an agreement that allows accessing the data for non commercial purposes. Data access will then be provided through an account in a secure cloud based environment.

## Code availability

The analysis code supporting this study is publicly available at https://github.com/hrossman/hpp-insulin-resistance-paper-public.

## Authors contribution

S.S conceived the project, designed and conducted the analyses, interpreted the results and wrote the manuscript. Y.T.B, M.G, D.A interpreted the results. A.G provided data, E.S provided data and interpreted the results, H.R conceived and directed the project and analyses, designed the analyses, interpreted the results, and wrote the manuscript.

## Declaration of generative AI and AI-assisted technologies in the manuscript preparation process

During the preparation of this work the author(s) used Claude (Anthropic) in order to assist with manuscript drafting, rephrasing and editing of text, verification of references, and refinement of terminology and classification frameworks. After using this tool/service, the author(s) reviewed and edited the content as needed and take(s) full responsibility for the content of the published article.

## Supplementary Figures

## Supplementary Figures

**Supplementary Figure S1.**
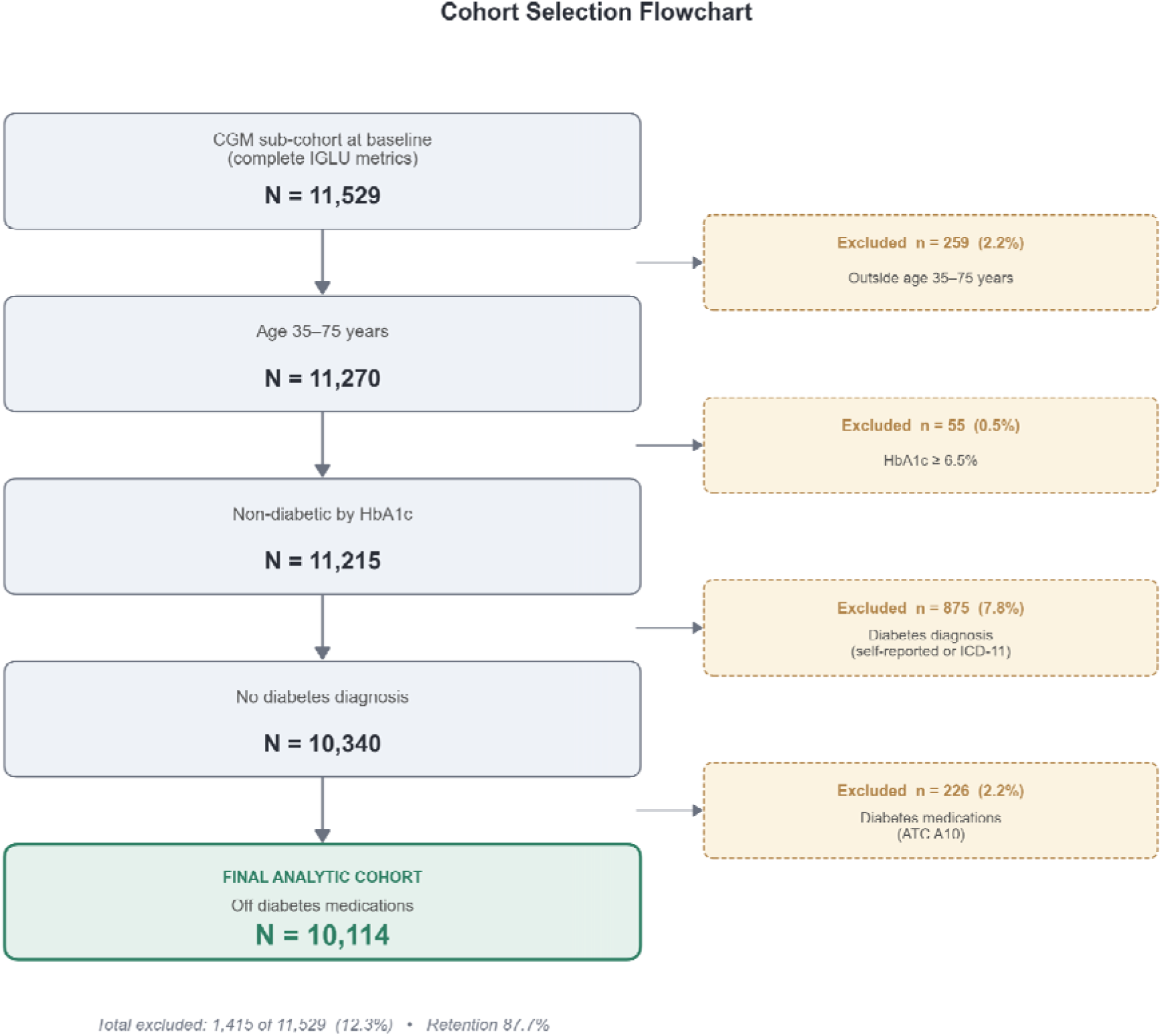
Cohort Selection Flowchart. flowchart showing sequential exclusion criteria from 11,529 CGM participants to the final cohort of 10,114 non-diabetic adults aged 35-75.

**Supplementary Figure S2.**
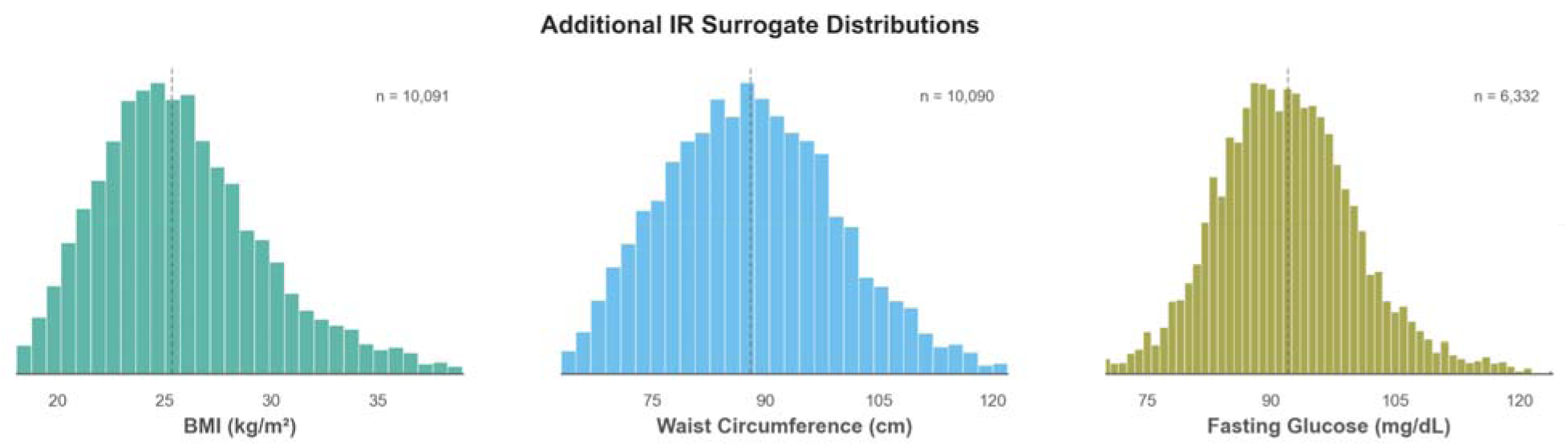
Distributions of BMI, waist circumference, and fasting glucose.

**Supplementary Figure S3.**
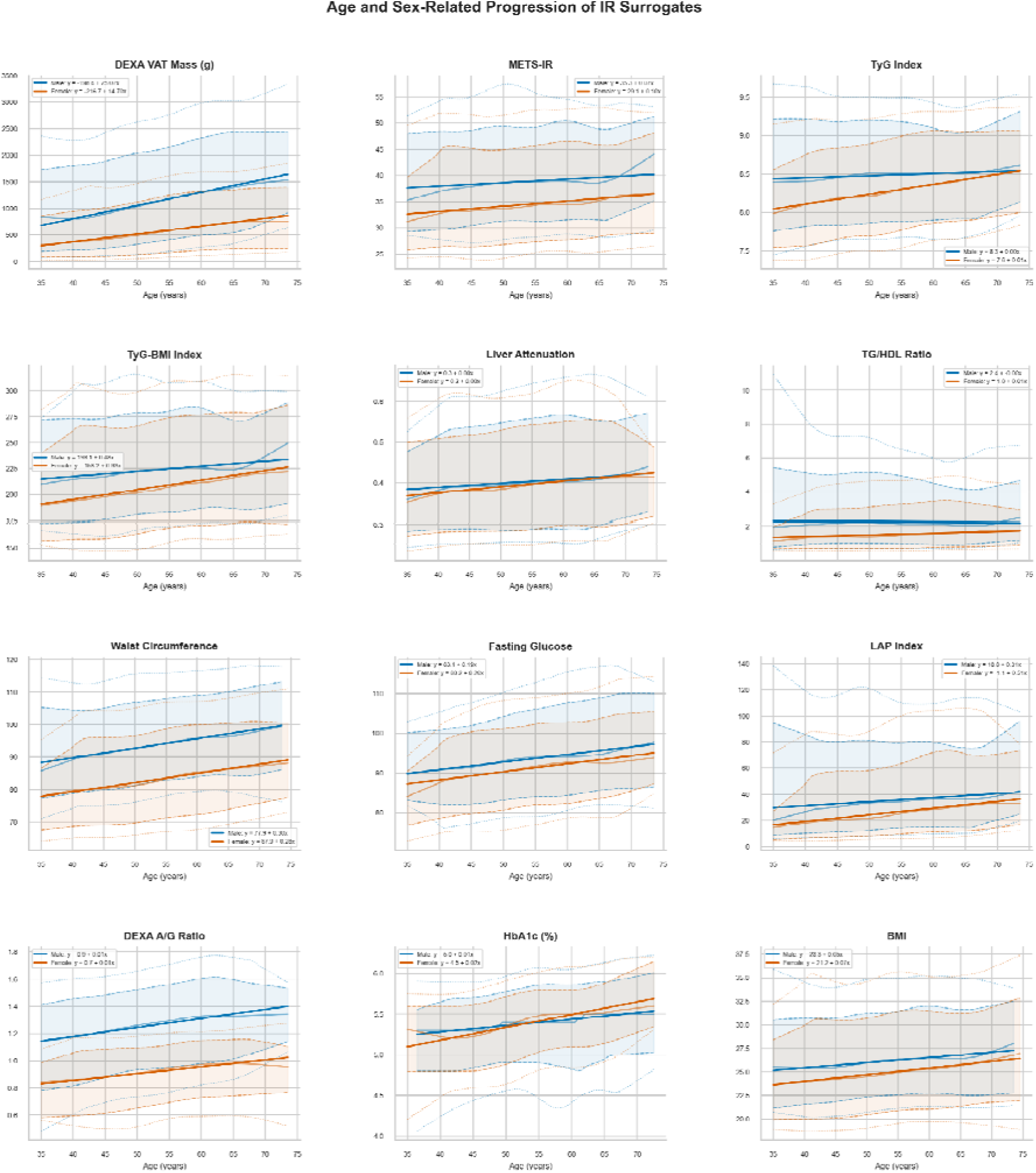
Age- and sex-specific progression of IR surrogate measures. LOWESS-smoothe percentile curves (3rd, 10th, 50th, 90th, 97th) with Huber robust regression equations shown by sex. Continuous age x-axis (35-75 years). Adiposity-based and lipid-derived surrogates show consistent age-related increases, with steeper male trajectories for VAT-based measures. Glycemic surrogates show more modest age effects.

**Supplementary Figure S4.**
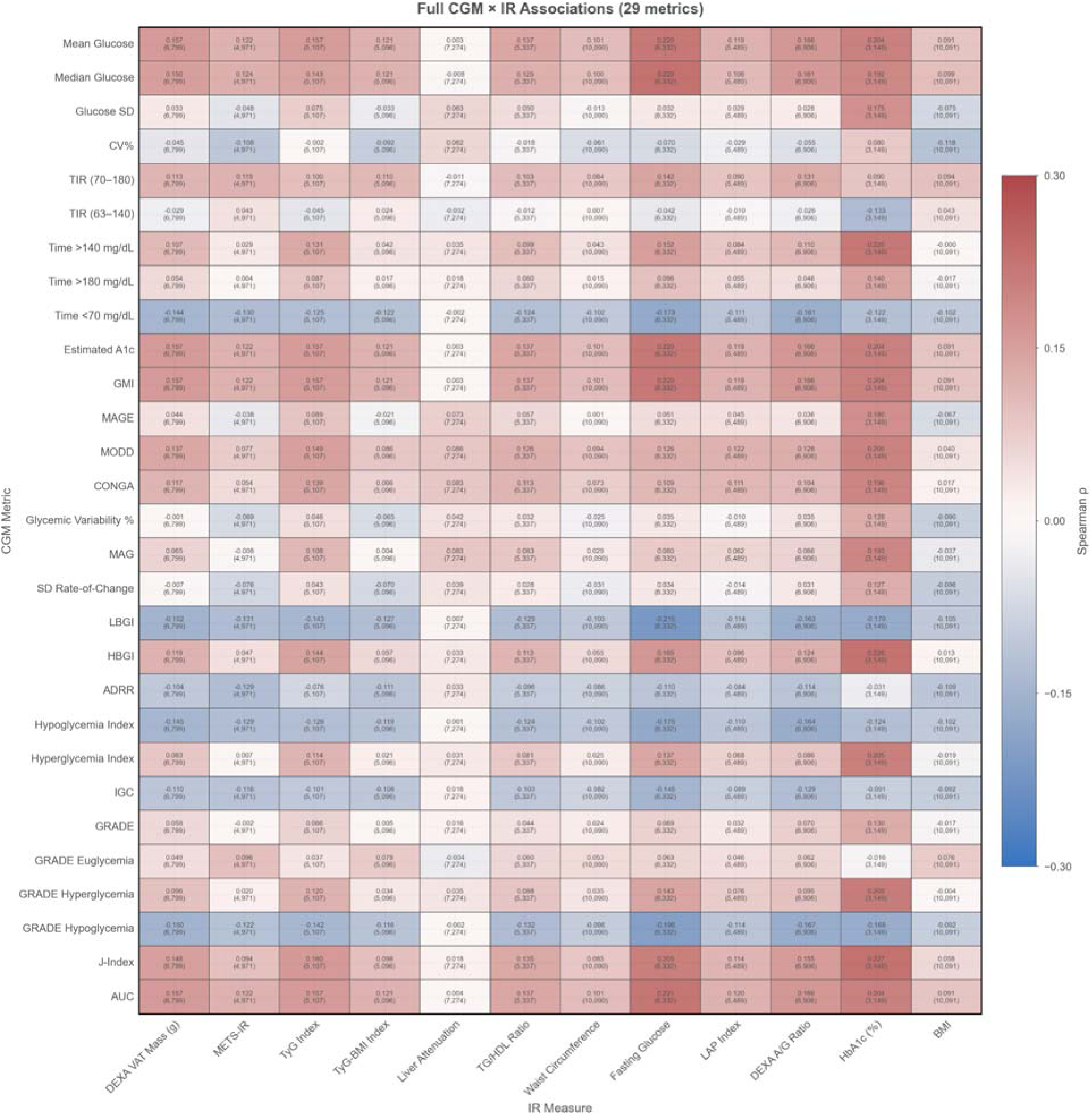
Full CGM-IR correlation heatmap showing all 29 IGLU-derived metrics and their Spearman correlations with IR surrogates. Cell annotations show rho and sample size (N). Extends Figure 2A to the complete set of CGM features available in both HPP and PPT cohorts.

**Supplementary Figure S5.**
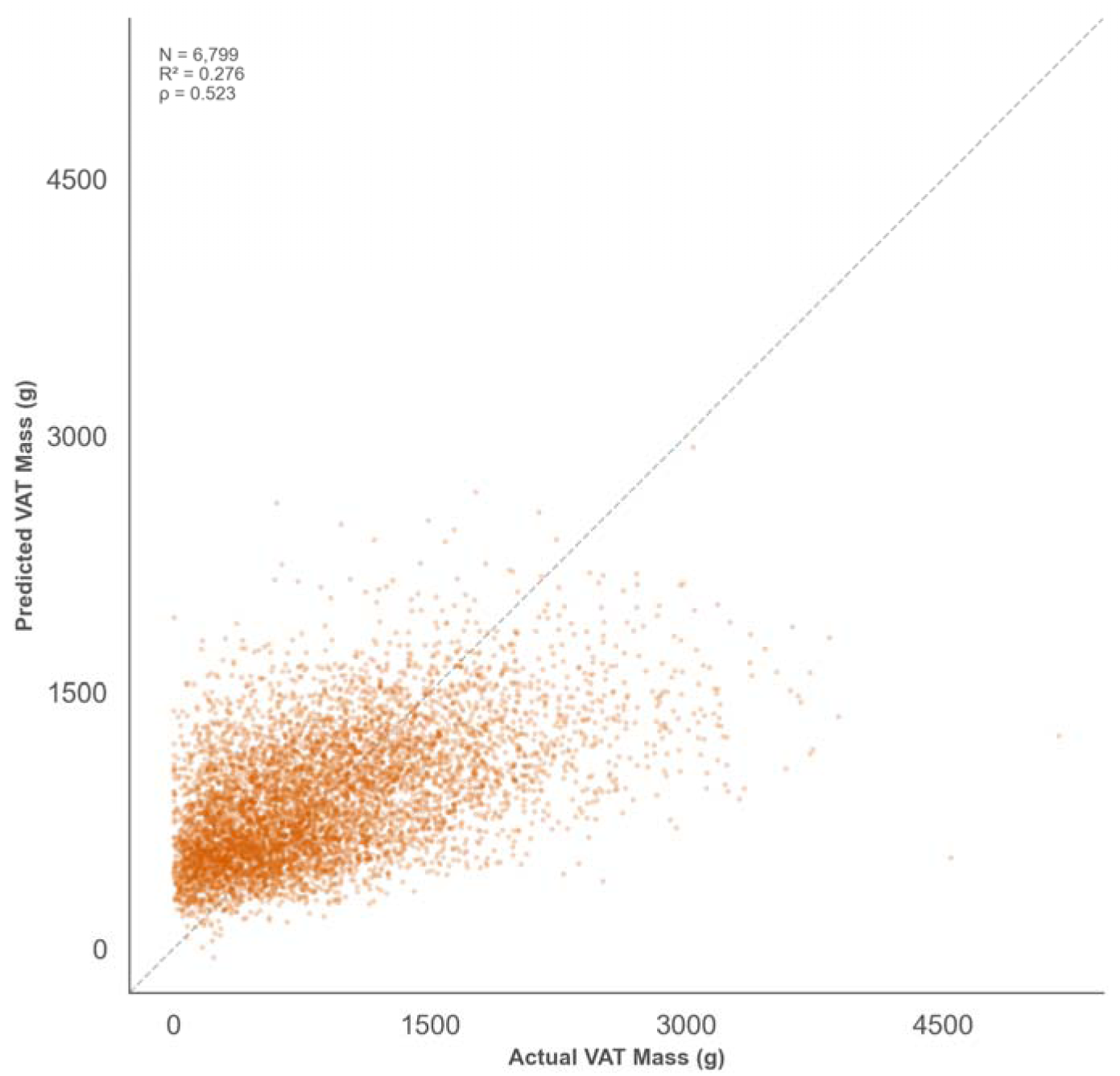
CGM-based prediction of VAT mass. Cross-validated predicted vs actual DEXA VAT mass using gradient boosting regression (N = 6,799; R^2^ = 0.276, Spearman rho = 0.524).

**Supplementary Figure S6.**
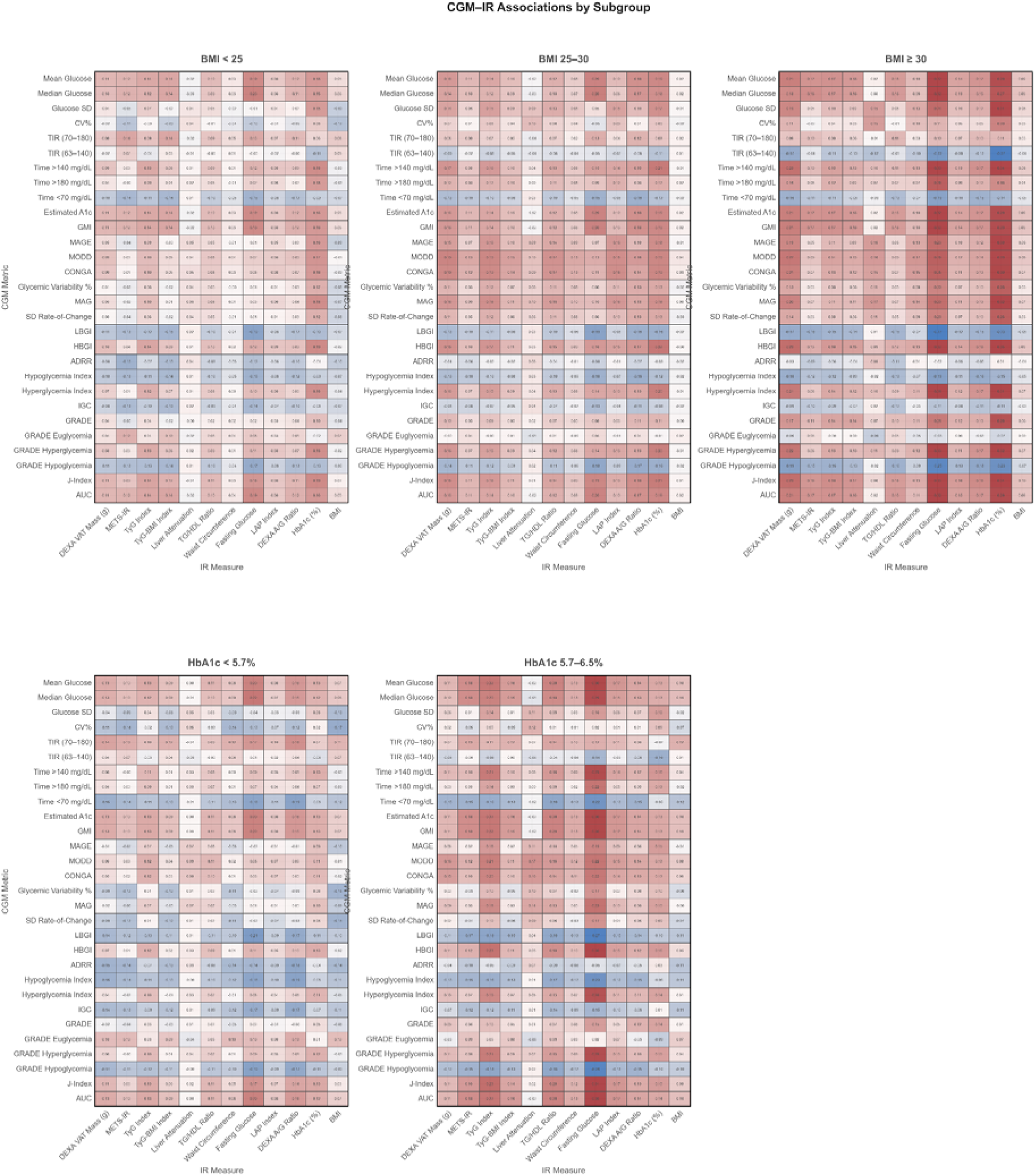
CGM-IR correlation heatmaps stratified by BMI and HbA1c showing all 29 IGLU-derived metrics. Computed within BMI (normal, overweight, obese) and HbA1c (normal, prediabetic) subgroups. CGM-IR associations strengthen with increasing metabolic burden, consistent with CGM metrics becoming more informative as underlying IR severity increases.

**Supplementary Figure S7.**
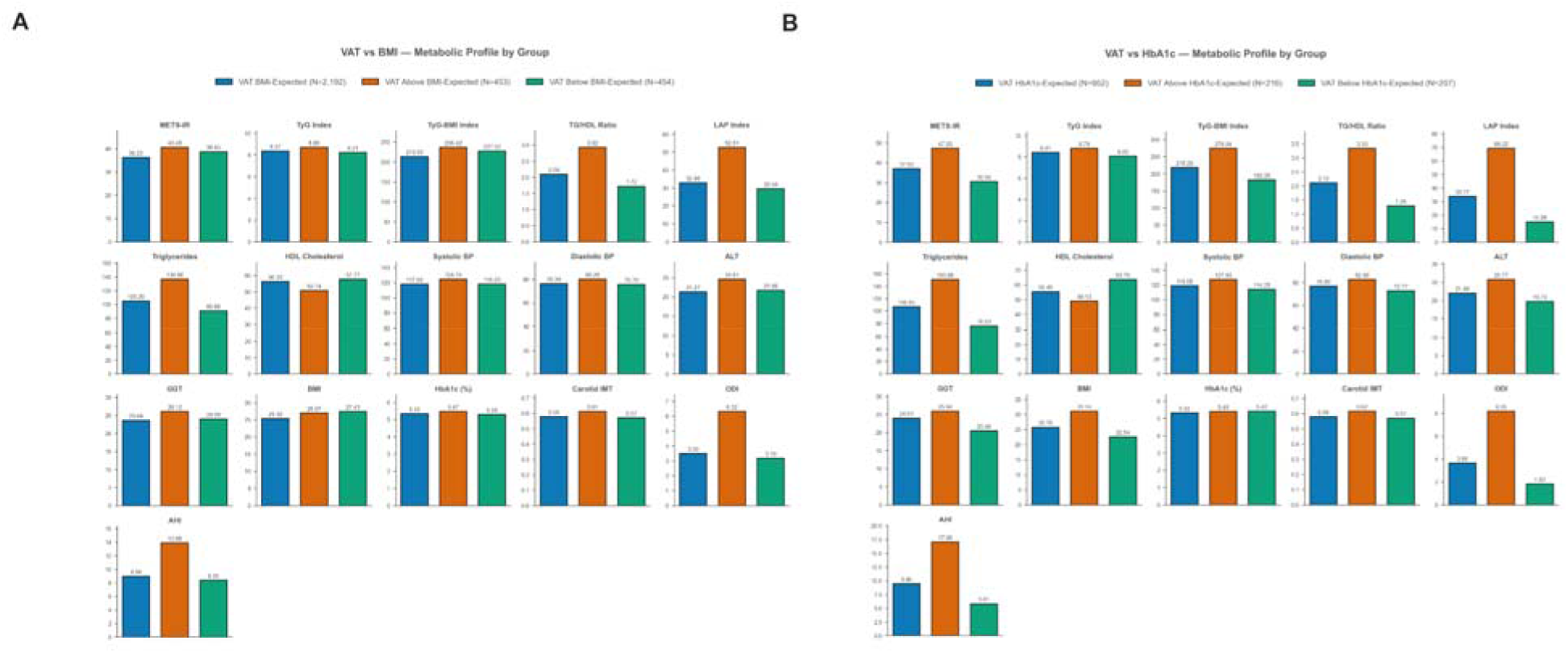
Discordance group metabolic profiles. Raw group-mean bar charts for clinical features across residual-based discordance groups on (A) the VAT-BMI axis and (B) the VAT-HbA1c axis. VAT Above BMI-Expected individuals show significantly worse metabolic profiles across all features examined; VAT Above HbA1c-Expected individuals show adverse lipid, hepatic, and adiposity profiles despite normal glycemi classification. Z-scored small-multiples forests are shown in Figure 3C-D. All between-group comparisons significant at FDR < 0.05 (Kruskal-Wallis test

**Supplementary Figure S8.**
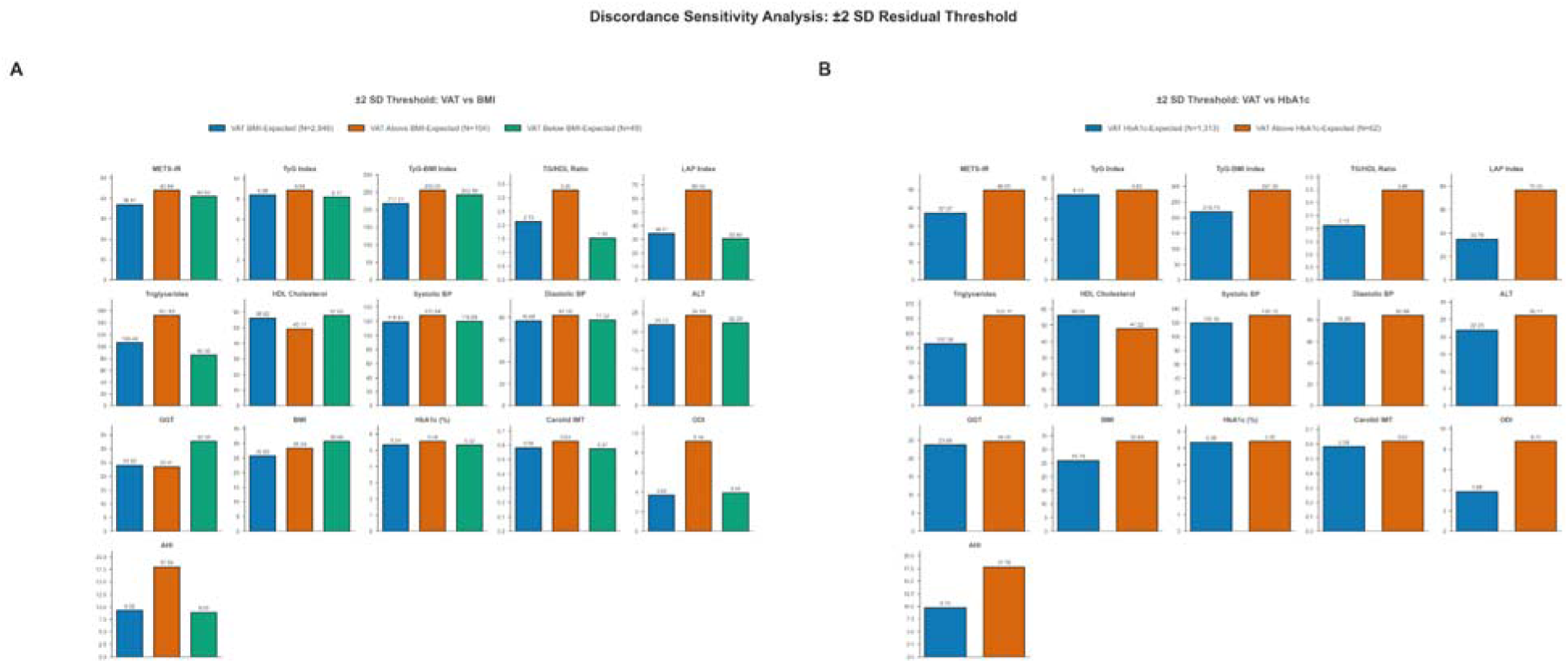
Discordance analysis using ±2 SD residual threshold showing that even when restricted to the residual extremes, metabolic profile differences between discordance groups in (A) the VAT-BMI axis and (B) the VAT-HbA1c axis and remain significant at FDR < 0.05, confirming that findings are robust t threshold choice.

**Supplementary Figure S9.**
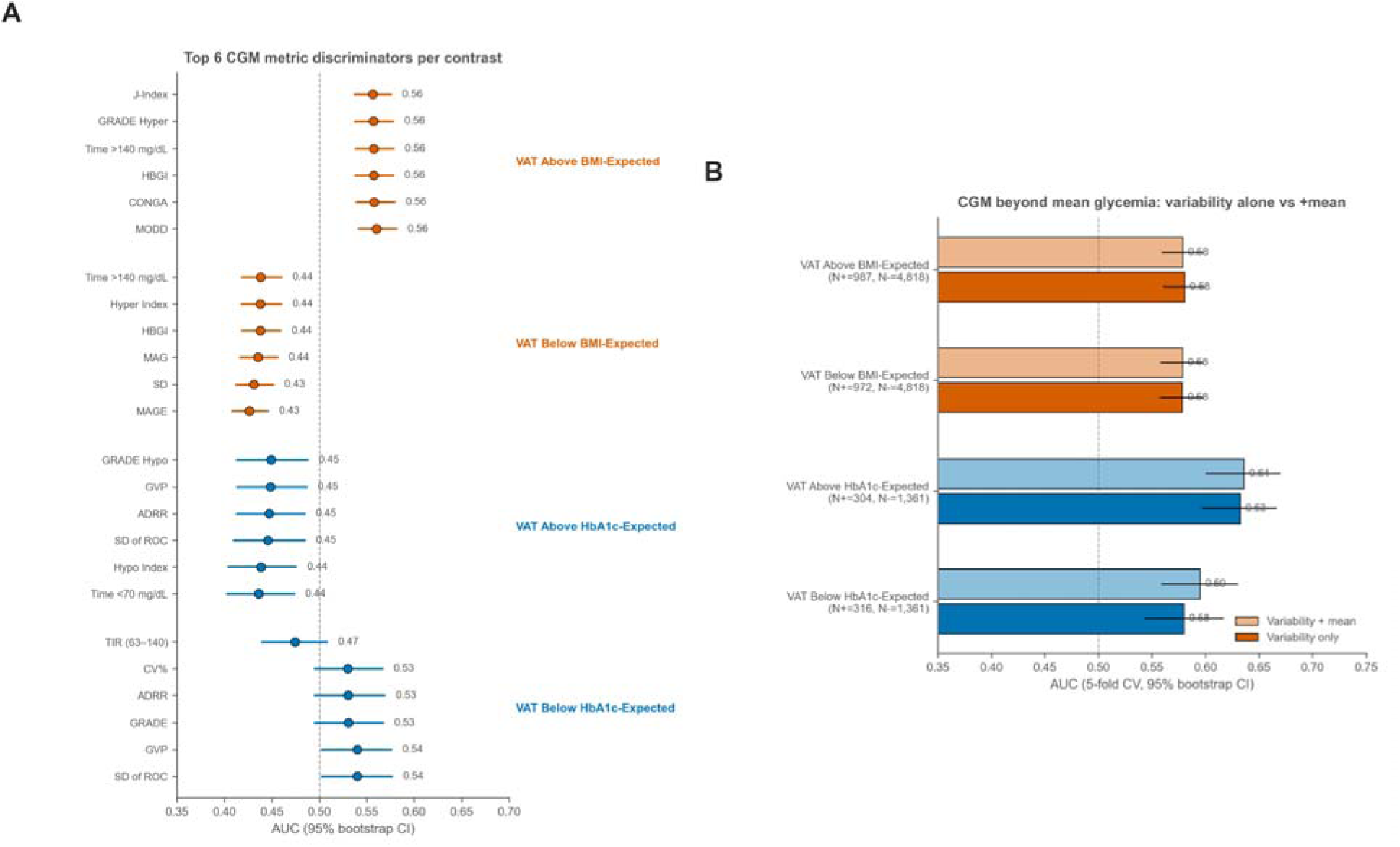
CGM discrimination of discordant VAT phenotypes. (A) Top six CGM metric discriminators per contrast - AUC with 95% bootstrap CIs (1000 iter, stratified resample); colour by axis (orange = VAT-BMI, blue = VAT-HbA1c); dashed line at AUC = 0.5. (B) Logistic-regression AUC for variability-only vs variability and glucose mean (5-fold CV, bootstrap 95% CI). For VAT Above HbA1c-Expected vs VAT HbA1c-Expected, adding mean glucose adds Delta AUC = +0.003.

